# Beckwith-Wiedemann syndrome and large offspring syndrome involve alterations in methylome, transcriptome, and chromatin configuration

**DOI:** 10.1101/2023.12.14.23299981

**Authors:** Yahan Li, Ping Xiao, Frimpong Boadu, Anna K. Goldkamp, Snehal Nirgude, Jianlin Cheng, Darren E. Hagen, Jennifer M. Kalish, Rocío Melissa Rivera

## Abstract

Beckwith-Wiedemann Syndrome (BWS) is the most common epigenetic overgrowth syndrome, caused by epigenetic alterations on chromosome 11p15. In ∼50% of patients with BWS, the imprinted region KvDMR1 (IC2) is hypomethylated. Nearly all children with BWS develop organ overgrowth and up to 28% develop cancer during childhood. The global epigenetic alterations beyond the 11p15 region in BWS are not currently known. Uncovering these alterations at the methylome, transcriptome, and chromatin architecture levels are necessary steps to improve the diagnosis and understanding of patients with BWS. Here we characterized the complete epigenetic profiles of BWS IC2 individuals together with the animal model of BWS, bovine large offspring syndrome (LOS). A novel finding of this research is the identification of two molecular subgroups of BWS IC2 individuals. Genome-wide alternations were detected for DNA methylation, transcript abundance, alternative splicing events of RNA, chromosome compartments, and topologically associating domains (TADs) in BWS and LOS, with shared alterations identified between species. Altered chromosome compartments and TADs were correlated with differentially expressed genes in BWS and LOS. Together, we highlight genes and genomic regions that have the potential to serve as targets for biomarker development to improve current molecular diagnostic methodologies for BWS.

## Introduction

Beckwith-Wiedemann Syndrome (BWS, OMIM #130650) is a congenital epigenetic disorder in humans with a spontaneous incidence of approximately 1 in 10,340 live births (1). The cardinal features of BWS include macroglossia, exomphalos, lateralized overgrowth, and hyperinsulinism (2). In addition, suggestive features of this syndrome include increased birth weight (>2 standard deviations above the mean), facial naevus simplex, ear creases/pits, transient hypoglycemia, and placentomegaly (2). BWS children have an increased likelihood of developing embryonal tumors most commonly Wilms tumor and hepatoblastoma (2). Molecular findings demonstrate an epigenetic origin to this condition with altered regulation of several imprinted domains foundational to its etiology. The most prevalent epimutation observed in this condition (∼50% of cases) is loss of DNA methylation (LOM) on the maternal allele at the imprinting center 2 (IC2; KvDMR1) which results in downregulation of the cell cycle regulator *CDKN1C* (2). The second most common epimutation (∼5% of cases) is gain of DNA methylation (GOM) on the maternal allele at the imprinted center 1 (IC1; *H19*/*IGF2*), which results in increased levels of *IGF2*, a potent fetal growth factor (2). In addition to these primary epimutations, BWS can also result from variation in DNA sequence at the above-mentioned imprinted domains with the most prevalent gene affected being *CDKN1C* (i.e. 5% of cases) (2).

The development of the mammalian embryo is a finely regulated process and adverse external insults during this window can have permanent influence on the health of the individual long after the exposure, a phenomenon known as Developmental Origins of Health and Disease (3, 4). An example of this phenomenon is seen in children conceived using assisted reproductive technologies (ART), which have over a 10-fold increased risk for developing BWS (∼1 in 1,126 live births) (5). The causal relationship between ART and the increased incidence of these syndromes is thought to occur by inducing errors in the epigenome of the embryos since ART manipulations overlap with critical developmental windows for epigenetic reprogramming (6–8).

Much of what is known to date regarding the etiology of the phenotypes of BWS has been achieved using mouse genetic models. For example, overexpression of *Igf2* resulted in macrosomia and disproportionate organ overgrowth (9), deletion of *Cdkn1c* causes abdominal wall defect (10, 11), and double mutation of *Cdkn1c* and *H19* with the adjacent 10 kb sequence (IC1 included) further results in macroglossia and placentomegaly (12). A shortfall of the mouse model, however, is that these genetic manipulations are often incompatible with fetal development and/or life after birth, and importantly, ART does not promote a BWS-like syndrome in mice. Large offspring syndrome (LOS) is a syndrome in bovine which phenocopies BWS (13). There are several important aspects of the bovine model that make this species well-suited to shed light into the complexities of the congenital human syndrome before and after birth. First, both species have a similar gestation length in which normally one offspring is gestated, and fetal landmarks occur at similar timepoints during pregnancy (14, 15).

This would prove valuable when trying to characterize the etiology and syndrome-related deviations in development, something that is not possible to characterize in mice. Second, BWS and LOS occur spontaneously, and ART promote their incidences (13, 16). Third, BWS and LOS share epigenetic defects in the form of loss-of-imprinting at IC2, downregulation of *CDKN1C*, and misregulation of several imprinted genes, including *PLAGL1*, *PEG3*, *IGF2R*, and several miRNAs in the *DLK1-DIO3* locus (17–23).

During the interphase of cell cycles, different chromatin loci interact to form functional structures including loops, topologically associating domains (TADs), compartments, and territories which are collectively referred to as chromosome architecture (24). Chromosome architecture can affect DNA methylation profile or be affected by DNA methylation changes (25, 26). Disruption of chromosome architecture may lead to aberrant expression of genes by gaining or losing interactions with active enhancers or changing their nuclear location and microenvironment, and can cause severe diseases in human (27). We and others have shown imprinted locus-specific alteration in chromosome architecture in these syndromes (28–30). A gap in knowledge that exists in the field, is the contribution that global architecture plays in BWS and the role that methylome play in this event. Our hypothesis is that genome-wide alteration of chromosome architecture occurs in BWS, which leads to regional and global gene expression changes. In this study, we systematically analyzed DNA methylation, chromosome architecture, and gene expression in fibroblast primary cells from 12 BWS patients with IC2 epimutation. With the advent of personalized medicine, the bovine model of BWS will be invaluable to be able to perform animal and *in vitro* studies in order to develop methodologies to correct epimutations, ameliorate the clinical features, and treat BWS. Therefore, we also performed similar analyses in LOS. This study represents the most comprehensive work done to date in BWS.

## Results and Discussion

### Genome-wide DNA methylation defects in BWS

Twelve BWS individuals with clinical molecular diagnosis of hypomethylation at IC2 were included in the study. LOM at CpG dinucleotides on the maternal allele at the imprinting control region (ICR) of *KCNQ1OT1*, (i.e. KvDMR1; also referred to as IC2), is the predominant epimutation in BWS, which accounts for ∼50% of documented cases. Whole genome bisulfite sequencing (WGBS) was conducted in fibroblast cells from the 12 BWS patients as well as from five controls to determine the status of DNA methylation (CpG) at base pair resolution for each group (Table S1). Results confirmed hypomethylation at IC2 (i.e. chr11:2719755-2722554) in BWS. The average DNA methylation at this locus was 17.59% in BWS samples and 45.18% in control samples (normal ∼50% since only the maternal allele is methylated at this ICR, within the range of the control samples). As expected, hypomethylation in BWS samples is not complete (i.e. 0%) because epimutations in BWS typically occur post-zygotically and are therefore usually mosaic which leads to a mixture of affected and unaffected cells being analyzed (2).

Methylome analyses identified 7,865 differentially methylated regions (DMRs) between BWS and control groups. Overall, both hypomethylated and hypermethylated DMRs in BWS are significantly enriched for gene promoters (1kb), gene bodies, CpG islands, CpG shores, CpG shelves, CTCF binding sites, and enhancers, and depleted for repetitive elements according to permutation tests (Figure S1A and Table S1). Among these examined genomic contexts, enhancers are the most enriched element overlapping DMRs in BWS (∼4X the incidence of random distribution).

DNA methylation level at gene promoter and first exon is negatively correlated with gene expression level (31, 32). For gene bodies, the DNA methylation and gene expression levels can affect each other. On one hand, transcription elongation recruits SETD2 to tri-methylate histone H3 lysine 36 which further recruits DNMT3A/B to methylate DNA and results in a non-monotonic relationship between gene expression and gene body DNA methylation levels (33). On the other hand, regulatory elements such as enhancers, genomic imprinting control regions, and CTCF binding sites overlap with gene bodies and the binding of methylation sensitive TFs can be affected by DNA methylation changes over these elements, therefore dysregulating the neighboring host genes (34, 35). In addition, DNA methylation level over exons can regulate alternative splicing events through genes such as CTCF, MECP2, and HP1 (36). CpG islands, CpG shores, and CpG shelves often overlap with promoters, enhancers, genomic imprinting control regions, or harbor CTCF binding sites (37). Repetitive elements usually contain high DNA methylation levels in order to be silenced for genome stability, however some repetitive elements or their transcripts can function as regulators for chromosome architecture and gene expression (38, 39). DNA methylation regulates gene expression mainly through affecting transcription factors (TFs) binding to their binding sites that contain CpGs (40). Binding of the insulator protein CTCF, is also regulated by this mechanism (40), when its binding motif includes CpGs (∼43% of CTCF binding sites) (41).

### Methylome distinguishes between clinically identified BWS IC2 individuals

Global DNA methylation analyses uncovered two clear sets of methylomes in the BWS population being studied (Figure 1 and S1A), with the main group composed of eight individuals (i.e. subgroup 1) and the other group composed of four individuals (i.e. subgroup 2). In addition, principal component analysis (PCA), similarly clustered the methylomes of the BWS individuals into two subgroups (Figure S2A). This finding was unexpected given that BWS individuals included in this study were all selected because they had low methylation levels at KvDMR1 in blood and therefore were clinically categorized as IC2 patients. Statistical comparisons of each subgroup and controls identified a larger number of DMRs in BWS subgroup 1 (n = 23,582) than in subgroup 2 (n = 1,915), most of which were hypermethylated (∼64% and 76%, respectively; Figure 1A and 1B). There were 672 DMRs identified as shared between the two BWS subgroups when compared to controls, with 97% of these DMRs having the same direction of methylation change (Figure 1C). For example, a hypomethylated region in both subgroups 1 and 2 (-62.31% and -17.46%, respectively) is within the gene body of *SKAP2* and overlaps an enhancer and predicted CTCF binding sites (Table S1). *SKAP2* is involved in the Src signaling pathway and regulates immune activation (42). Altered DNA methylation at this locus, is associated with significant downregulation of this transcript (4.7X) in BWS subgroup 1 (Table S2). An example of a shared hypermethylated DMR (19.68% and 15.77%, respectively) resides in the gene body of *AMZ1* which encodes a metallopeptidase that is predicted to be involved in proteolysis (43). This DMR also overlaps an enhancer and predicted CTCF binding sites and associates with a significant downregulation (2.7X) of this gene in BWS subgroup 1.

**Figure 1.**
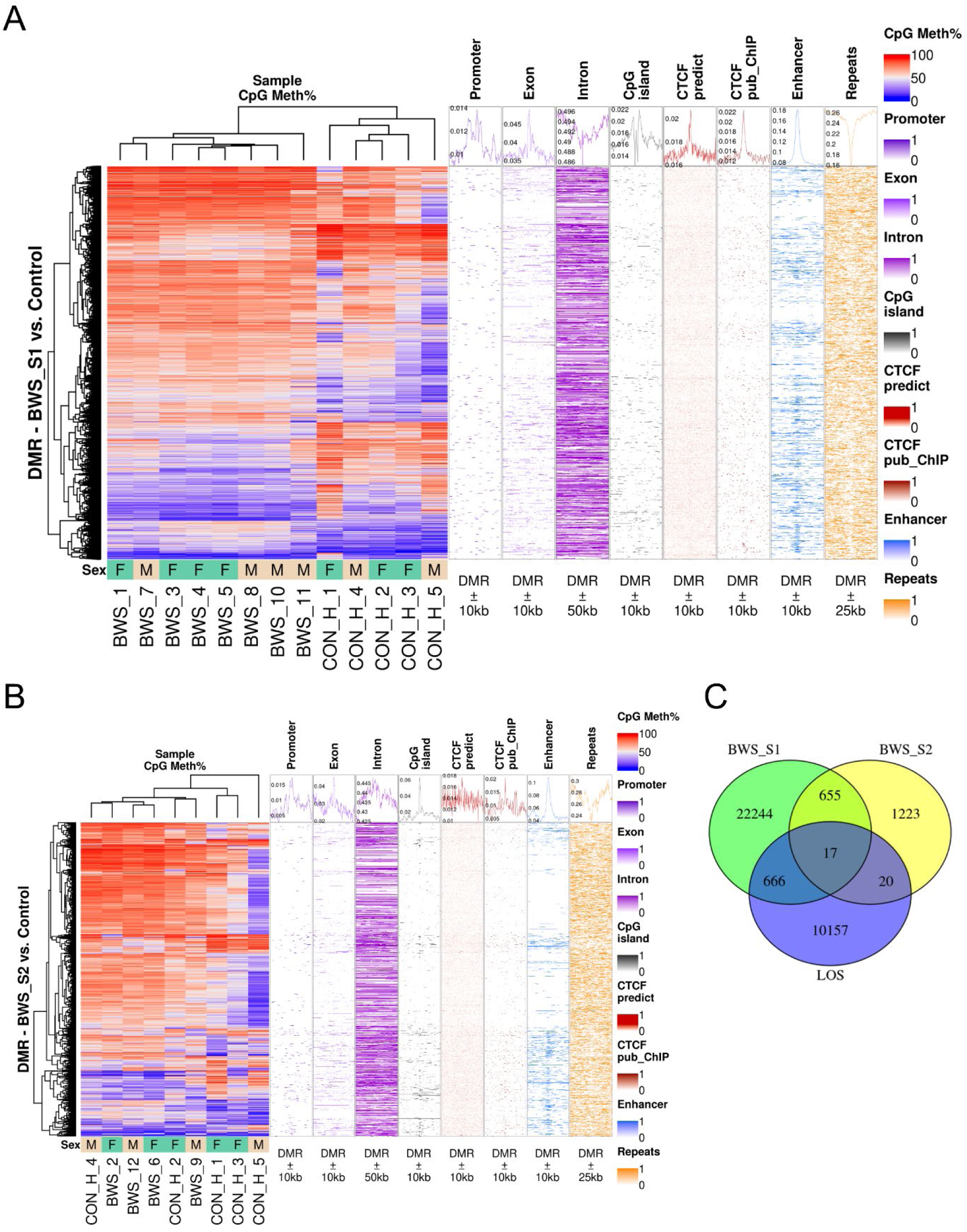
CpG methylation heatmap and genomic context enrichment for differentially methylated regions (DMR). **(A)** and **(B)** DMR for human BWS subgroup 1 compared with control group **(A)**, and human BWS subgroup 2 compared with control group **(B)**. The left panel is the heatmap with hierarchical clustering and sex annotation which include male (M) and female (F). The right panels are the distribution of genomic contexts at the DMR (centered) and surrounding regions. Genomic contexts include 1kb promoter, exon, intron, CpG island, predicted CTCF binding sites, public CTCF ChIP-seq peaks, enhancers, and repetitive sequences. The line chart on top of each panel is the average enrichment score for all the DMRs. **(C)** Number of unique and overlapped DMRs between BWS subgroups and LOS.

Interestingly, although these examples show congruency of DNA methylation epimutation, the fact that subgroup 2 does not mimic the altered gene expression observed in subgroup 1 indicate that additional mechanisms of gene regulation are at play.

Among the top 50 more severely epigenetically dysregulated regions (both hyper- and hypomethylated) in the BWS subgroup 1, 12 are located within the *HOXA-D* clusters (Figure 2 and Table S1). Homeobox genes are evolutionary conserved transcription factors that control morphogenetic programs during development. The 39 transcription factors are located in four gene clusters denoted *HOXA*, *HOXB*, *HOXC*, and *HOXD*. Dysregulation and genetic mutations of *HOX* genes has been observed in a sundry of cancers and developmental abnormalities (44). In addition, both hyper- and hypomethylated *HOX* genes are frequently observed in cancer, including Wilms tumor, a commonly diagnosed tumor in BWS (45). In total, there are 66 DMRs in BWS subgroup 1, a dysregulation not observed in BWS subgroup 2, which only has 5 DMRs within the four *HOX* gene clusters. Shared hypomethylated DMRs between subgroup 1 and 2 are located within the *HOXA3*, *HOXB6*, and *HOXB7* genes while the DMRs at the *HOXB3* has opposite erroneous DNA methylation levels between the two subgroups. An interesting observation of our study is that regions of hypermethylation and hypomethylation alternate within the *HOX* gene clusters (including *HOXA*) in BWS subgroup 1 suggesting that altered chromatin topology at these loci may be involved in this syndrome (Figure 2; see Hi-C section). *HOXA1*, *HOXA5*, *HOXA6*, are downregulated in BWS subgroup 1. Erroneous expression of *HOXA1* has been reported in ear malformations in humans and pigs (46, 47). Altogether, identified DMRs in BWS reflect altered *HOX* expression during embryonic development and have the potential to serve as molecular markers to distinguish between BWS IC2 subgroup 1 and BWS IC2 subgroup 2.

**Figure 2.**
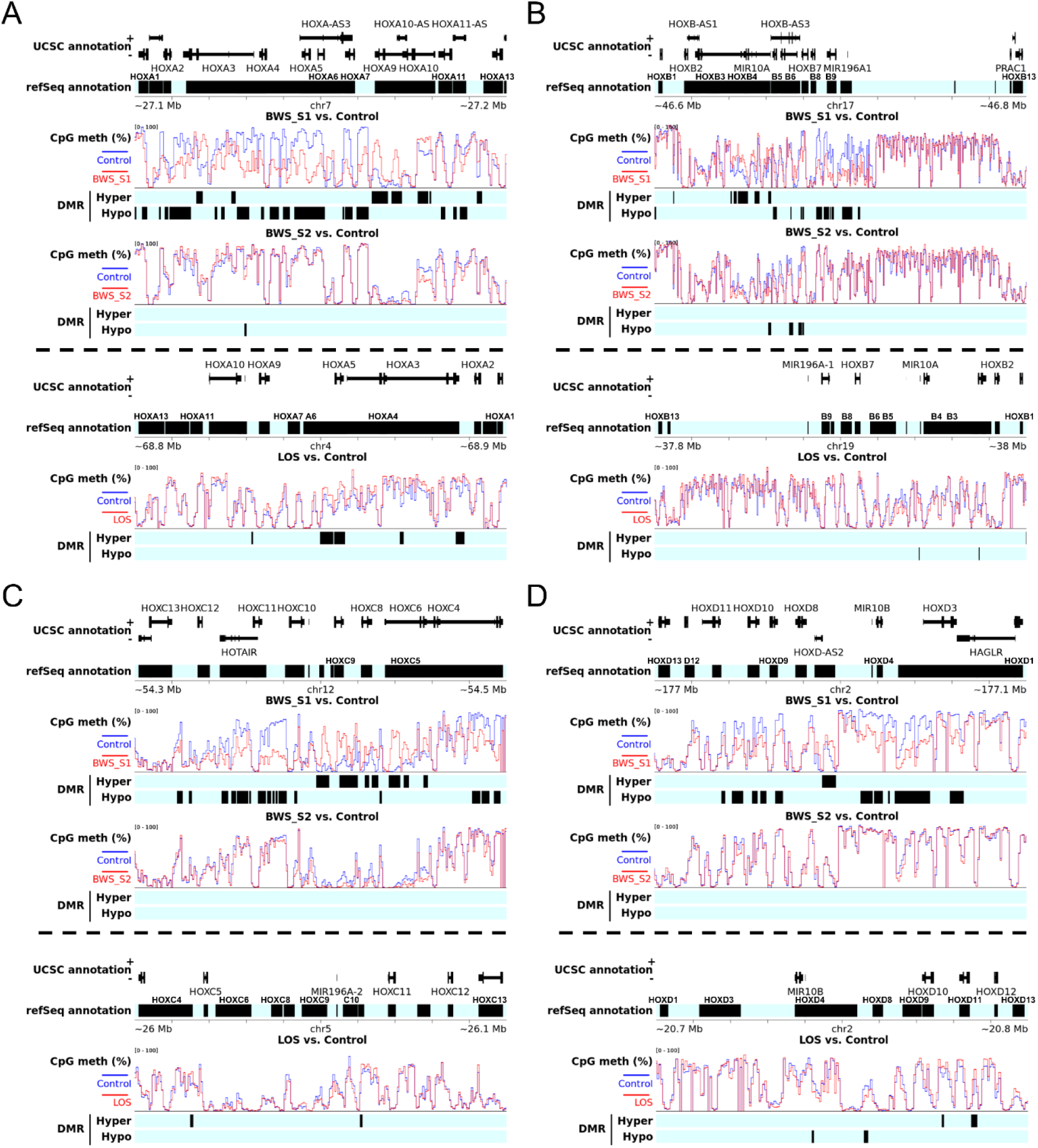
DNA methylation alterations at *HOX* gene clusters in BWS and LOS. **(A)** *HOXA* cluster. **(B)** *HOXB*. **(C)** *HOXC*. **(D)** *HOXD*. Top panel is human genome and bottom panel is bovine genome for each plot. Additional gene annotation from NCBI RefSeq is added since some *HOX* genes are not annotated in the R package for UCSC annotation. All the *HOX* genes from the same cluster are located on the same strand. The order of *HOX* genes may be opposite for human and bovine.

**Figure 3.**
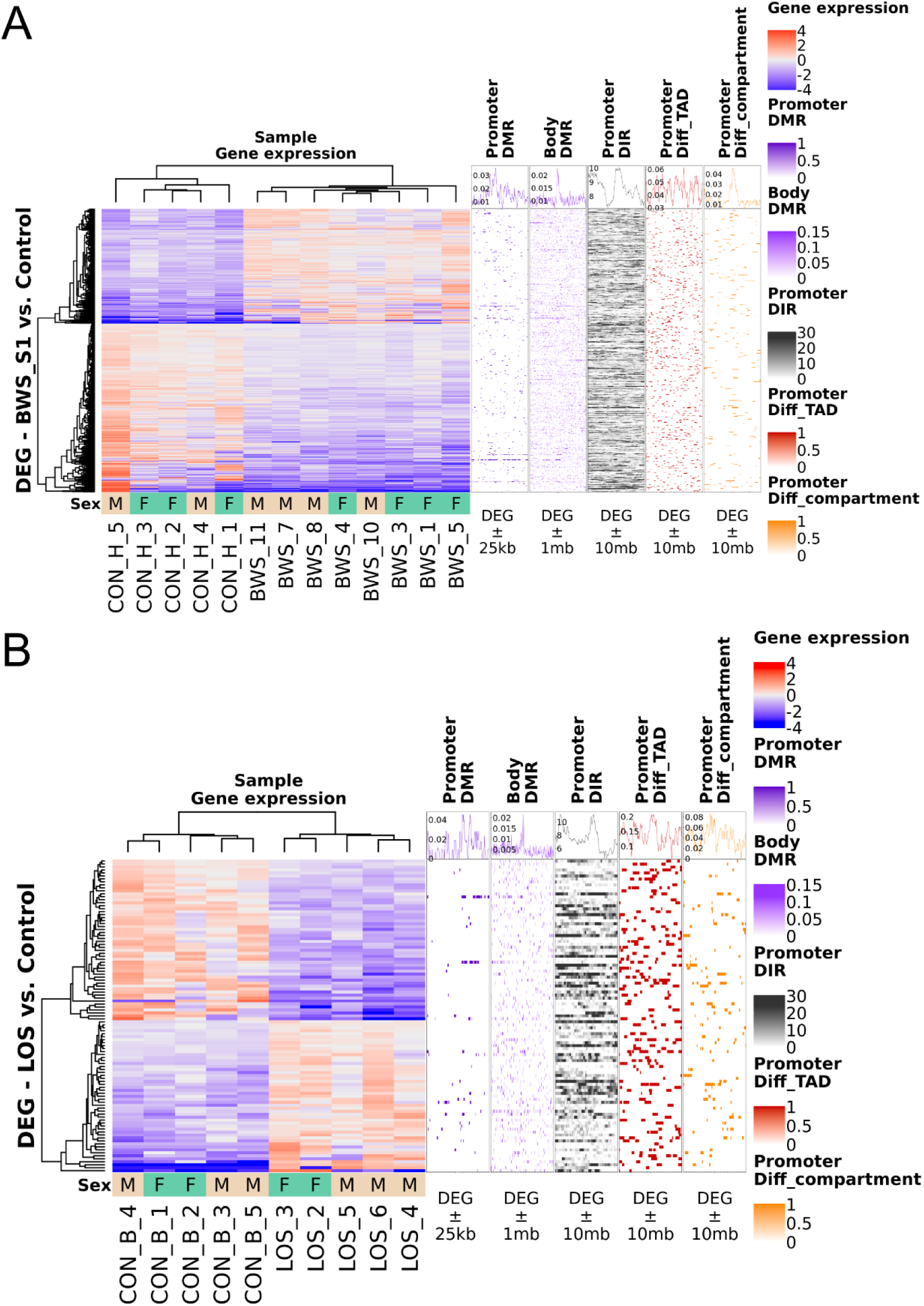
Expression heatmap and genomic context enrichment for differentially expressed gene (DEG). DEG for human BWS subgroup 1 compared with control group **(A)**, and bovine LOS group compared with control group **(B)**. The left panel is the heatmap with hierarchical clustering and sex annotation which include male (M) and female (F). Gene expression is shown as log2 transformed difference between sample and row average. The right panels are the distribution of genomic contexts at the DEG (centered at 1kb promoter or gene body) and surrounding regions. Genomic contexts include differentially methylated regions (DMR), differential interacting regions (DIR), differential topologically associating domains (diff_TAD), and differential compartments identified in corresponding group comparisons. The line chart on top of each panel is the average enrichment score for all the DEGs.

### Conservation of DNA methylation defects between BWS and LOS

We performed similar analyses in the bovine BWS model (LOS) to those mentioned above (Figure S2, S3, and Table S1). Here, we will focus our discussion on similarities in methylation errors between LOS and control as those observed between BWS and control (all results are available in the supplementary materials). Similar to BWS, the majority (75%) of DMRs were hypermethylated in LOS (Figure S3A). We then compared DMRs between LOS and each of the two BWS subgroups and found that 683 DMRs were shared with subgroup 1 with 74% changed in the same direction, and 36 DMRs were shared with subgroup 2 with 53% changed in the same direction (Figure 1C and Table S1).

A shared hypomethylated DMR in BWS subgroup 1 and LOS resides in the gene body of *BCL11A* (-29.85% and -15.42%, respectively) and overlaps with an enhancer. *BCL11A* is a transcriptional repressor which regulates blood cell development and is significantly upregulated 3.6X in LOS and shows a trend towards upregulation (2.4X, FDR = 0.056) in BWS subgroup 1 (Table S2). An example of a shared hypermethylated DMR between BWS subgroup 1 and LOS is found in the gene body of *PTPRE* (27.35% and 18.35% increased DNA methylation respectively). This DMR also overlaps with an enhancer. *PTPRE* encodes a receptor protein tyrosine phosphatase and plays an essential role in cell signaling and is significantly downregulated (2.9X) in BWS subgroup 1. In addition, DNA hypermethylation was observed at *HOXA3*, *A5*, *A6*, *C4*, *C10*, and *D11*, and hypomethylation was observed at *HOXB3* and *D4* in LOS. Shared DMRs between BWS subgroup 2 and LOS that overlap enhancers include hypomethylation at the gene body of *LDLRAD3*, *ZNF536*, and hypermethylation within *KLHL26* and *EBF3*.

More importantly, these multispecies comparison analyses allowed us to identify 17 DMRs that are shared between both BWS subgroups and the bovine animal model. These included the three mentioned above for subgroup 2 (except *ZNF536*), hypomethylation at the promoters of *KCNQ1OT1* (i.e., KvDMR1; IC2) and in the gene bodies of the epigenetic modifiers *HDAC4* (which also overlaps an enhancer) and *PRDM8* (Figure 1C and Table S1). Further, another shared DMR resides in the gene body of *HOXA3* and overlaps with CTCF binding sites and an enhancer and is hypomethylated in both BWS subgroups and hypermethylated in LOS, which suggest this to be a labile locus. Gene body DNA methylation is associated with active transcription and altered DNA methylation level in this context would indicate that at the time of collection, and/or previously, the genes of these histone modifiers were differentially expressed between affected individuals and controls. Further, one of the shared hypomethylated DMRs is in the gene body of *NR2F1-AS1*, a gene associated with hepatocellular carcinoma (48, 49). These 17 genomic regions should be further analyzed in future studies to determine their diagnostic usefulness in the clinic.

### DNA methylation defects at imprinted domains in BWS and LOS

Genomic imprinting is an epigenetic mechanism that orchestrates the parental allele-specific expression of a subset of mammalian genes which control growth and development of the conceptus. As mentioned previously, BWS is considered a loss-of-imprinting condition, therefore, we next assessed the level of methylation at all known and candidate imprinted loci in humans. We only identified IC2 hypomethylation (-27.6%) when all BWS were compared to controls (Table S1), an expected finding as hypomethylation at this ICR was the criterion used to select patients to include in this study. For BWS subgroup 1, in addition to IC2 (-28.4%), hypomethylation at *MEG3* TSS-DMR (-15.2%) and putative ICR of *JAKMIP1* (-15.9%), and hypermethylation at *H19* gene body (17.2%) were detected. *MEG3* is a long non-coding RNA with tumor suppressor roles within the *DLK1*-*DIO3* imprinted cluster, and hypomethylation at *MEG3* TSS-DMR is a molecular marker of Temple Syndrome and has also been reported with BWS patients (50). For BWS subgroup 2, in addition to IC2 (-26.2%), hypomethylation at *GNAS* ICR (*GNASXL*, -16.7%) and over the third CTCF binding site (51) of IC1 (-17.5%) were detected. These findings indicate the involvement of multi-locus imprinting disturbance (MLID) in the BWS samples. MLID refers to a condition that patients with imprinting disorders have DNA methylation changes at multiple imprinted loci across the genome in addition to the primary lesion (52). MLID occurs more frequently in imprinting disorders caused by LOM at key imprinted loci than GOM (e.g. BWS with IC1 GOM), and about one-third of BWS IC2 LOM patients have MLID (2). The presence of MLID suggests abnormal epigenetic reprogramming during oocyte and/or early embryo development for these BWS patients (52).

Similar to BWS, LOS group showed hypomethylation at IC2 (-19.4%). In addition, hypomethylation was observed at ICRs of *PEG10* (-19.0%), *NAP1L5* (-16.5%), *IGF2R* (-26.7%), *GNAS* (*GNASXL*, -15.6% to -19.2%), *NNAT* (-16.1% to -18.9%), *IGF1R* (-21.5%), and *INPP5F* (-20.2%), while hypermethylation was detected at the ICR of *GNAS* (first exon and intron, 16.4%). It should be noted that the difference observed in the level of methylation between LOS and BWS is likely due to the fact that the human samples were chosen based on hypomethylation of IC2 while the LOS were not.

Since it is not feasible to perform parental allele-specific analyses with the human samples, we employed the bovine BWS model for these determinations. Allele-specific methylation analysis in the control group clearly separates the maternal and paternal alleles in the PCA plot (Figure S2C) and identified 10,143 DMRs between the parental alleles, with the majority (>99%) being hypomethylated on the paternal allele (Figure S3C). Twenty-four of the identified DMRs overlap known imprinted genes and exhibit the expected allelic patterns, including *PEG10*, *MEST*, *NAP1L5*, *PLAGL1*, *IGF2R*, *KBTBD6*, *GNAS (*both promoter and *GNASXL)*, *PEG3*, *MAGEL2*, *SNRPN*, *IGF1R*, *INPP5F*, *KCNQ1*, and *H19* (Table S1). Allele-specific comparisons between LOS and controls, identified that both alleles are mainly hypermethylated in LOS, which is similar to the finding of the overall methylome comparisons in BWS and LOS (Figure S3D and S3E). This indicates that genome-wide gain of methylation seems to be more prevalent than loss of methylation in the syndrome. Maternal allele hypomethylation in LOS was found at the ICR of *NAP1L5* (-30.4%), *GNAS* (*GNASXL*, -36.3%), *MAGEL2* (-35.9%), *SNRPN* (-20.1% to -29.4%), *IGF1R* (-54.9%), and *INPP5F* (-42.3%), and maternal allele hypermethylation at *GNAS* ICR (promoter and first exon and intron, 15.8% to 44.1%). As expected, the normally methylated maternal allele of the *IGF2R* ICR is hypomethylated (-66.41%) in LOS when compared to controls, similar to what has been previously published by us and others (19, 53).

The range of methylation for the maternal allele at IC2 is 84.6% to 94.5% in controls and 36.8% to 93.4% in LOS samples. However, the group mean difference between LOS and control is 14.7% at the maternal allele of IC2, which does not pass the 15% filtering threshold and therefore was not included in the results. For IC1, normal ranges of allele-specific methylation were observed in both control and LOS groups, which is similar in human that the bulk methylation level at this locus had no change in BWS. The paternal allele of *H19* gene body exhibits higher average methylation level (55.2%) when compared to the maternal allele (9.7%) in bovine controls. Although the IC1 methylation is not widely changed in either BWS or LOS, we detected hypermethylation in the gene body of *H19* in BWS subgroup 1 (17.2%) and LOS (15.8%) at the bulk level analyses, and on the paternal allele in LOS (21.0%), which is a novel finding of yet unknown significance. As we learn more of the signatures in BWS, we will be able to better select bovine samples that more closely mimic the epimutations of each human subgroup in molecular studies.

In summary, the abovementioned findings demonstrate methylome distinctions between BWS samples within the same molecular diagnosis of IC2 hypomethylation, and the two BWS subgroups show unique MLID patterns. BWS subgroup 2 has fewer methylation epimutations than subgroup 1 and even clusters with the controls in the unsupervised analyses. This is a novel finding, and future analyses will need to determine if this is a consistent observation for the BWS IC2 population at large, which would help clinicians make more informed treatment decisions.

### Integrating transcriptome and methylome analyses reveals regulated gene expression pattern

Similar to the DNA methylation findings, the transcriptomes of the BWS samples separated into two subgroups with the same composition of samples in the PCA pot (Figure S2D). As before, BWS subgroup 2 samples were intermixed with the controls, which indicates that samples in subgroup 2 have fewer molecular deviations than those in subgroup 1 when compared to the controls. Only eight differentially expressed genes (DEGs) were detected when all BWS samples were compared to controls (Table S2). Of those *MAS1*, *LINC01234*, *PECAM1* were upregulated and *FAM225A*, *LINGO2*, *MIR12121*, *MIR92A1*, and a novel transcript were downregulated.

For the subgroup specific analyses, altered transcript levels were identified in 742 genes in BWS subgroup 1 and two genes in subgroup 2 when compared to controls. For subgroup 1, the top five upregulated genes were *CXCL14*, *PECAM1*, *GRPR*, *NTRK3*, and *COL23A1*, and the top 5 downregulated were *HOXA6*, *IFIT2*, *APBB2*, *OSBPL10*, and *SLC14A1*. *MAS1* is also upregulated in BWS subgroup 1, and is a gene which we have found in bovine to be normally incorporated in a topological associating domain (TAD) which contains the *IGF2R* ICR (*AIRN* promoter) in the unmethylated paternal allele but not on the methylated maternal allele (30). In LOS, hypomethylation at this ICR on the maternal allele permits binding of the insulator protein CTCF and the formation of a TAD which erroneously encompass the *MAS1* gene. Different from LOS, *IGF2R* ICR hypomethylation was not detected in BWS, which is expected since *IGF2R* is generally considered not imprinted in human (54). However, *IGF2R* showed a trend of downregulation (1.7X, FDR = 0.058) in BWS subgroup 1 which is similar to LOS, and could be associated with the *MAS1* misregulation. IGF2R protein functions as a scavenger receptor for IGF2 to regulate its degradation, and IGF2R protein downregulation and malfunctioning (retained in cytoplasm) has been reported in BWS (55). Further many DEGs of subgroup 1 are associated with tumorigenesis, including downregulation of *MDGA2*, *NGFR*, *BMP2*, and upregulation of *PECAM1* and *SESN3* which is also involved in regulating insulin sensitivity (56–61). In addition, four DEGs are associated with DMRs at their promoters including upregulated *KCNQ1OT1*, *CLEC14A*, and *SOST* which exhibited hypomethylation at their promoters and *TGM2* which is downregulated and experienced promoter hypermethylation. The two genes with altered expression in BWS subgroup 2 are the silenced *PRKAB2* and an upregulated novel transcript and were not observed as dysregulated in subgroup 1. *PRKAB2* encodes a regulatory subunit of the AMPK which both up- and downregulation has been observed in various cancers including liver cancer (62).

KvDMR1, also referred to as IC2, is the promoter of the paternally expressed gene *KCNQ1OT1*. This lncRNA acts in cis by bringing Polycomb group protein-induced chromatin silencing to the flanking imprinted genes in the locus (63), including the cell-cycle regulator *CDKN1C*. Loss of methylation on the maternal allele at IC2 results in increased transcript amount of *KCNQ1OT1*. On average, BWS subgroup 1 had a ∼3.4 fold increased expression of this gene (FDR = 0.003), and *CDKN1C* showed an ∼50% decrease (not significant, FDR = 0.47). The expression of these gene in BWS subgroup 2 were similar to that of the controls. For all BWS samples, the correlation coefficient (r) between IC2 methylation level and expression of *KCNQ1OT1* and *CDKN1C* are -0.37 (p = 0.24 with a two-tailed t-test) and 0.71 (p = 0.01), respectively. For subgroup 1 samples, r for *KCNQ1OT1* and *CDKN1C* are -0.72 (p = 0.044) and 0.42 (p = 0.3), and for subgroup 2 samples, r are -0.04 (p = 0.96) and 0.95 (p = 0.046), respectively.

Expression of genes require the action of specific transcription factors as well as interactions with the enhancer. The results that hypomethylation at IC2 does not result in increased expression of *KCNQ1OT1* in subgroup 2 indicates that upstream events are also affected differentially between these subgroups. Regarding the association between changes in gene expression and DNA methylation, upregulated DEGs in BWS subgroup 1 showed significant enrichment for hypermethylated DMRs at promoter and hypomethylated DMRs at gene body, and downregulated DEGs showed enrichment for both hyper- and hypomethylated DMRs at gene body according to permutation tests (Table S2).

Alternative splicing (AS) is a molecular mechanism that regulates the generation of multiple (m)RNA and protein products from a single gene. Basic events include exon skipping, alternative 5′-splice site, alternative 3′-splice site, mutually exclusive exons, and intron retention (64). Given that DMRs overlapping exons can influence alternative splicing of pre-(m)RNA, we analyzed the total RNA sequencing data to identify AS events and their putative relationship to DNA methylation. In total 2230 AS events were identified in BWS subgroup 1, and 1140 in BWS subgroup 2 when compared to the control group (Figure 4A and Table S2). For subgroup 1, the host genes of 39 AS events are also DEGs (n = 27), including genes with important fetal development roles such as *BCL2L1*, *CDKN1A*, *DUSP6*, *GSDME*, *SEMA3C*, *TGFBR2*, and *ZMAT3*, and the exons of 73 AS events overlap with DMRs. For subgroup 2, none of the AS events is associated with DEGs or DMRs. There were 646 AS events shared between the two BWS subgroups, including *MTOR*, *IGF1R*, *MEG3*, *SETD5*, *HMGA1*, *CDKAL1*, and many more genes involved in fetal development and tumorigenesis. AS events appeared to be evenly distributed across chromosomes, and most were skipped exons with an average of 43.1% across pairwise comparisons. These data suggest alternative splicing as a contributor in the etiology of BWS.

**Figure 4.**
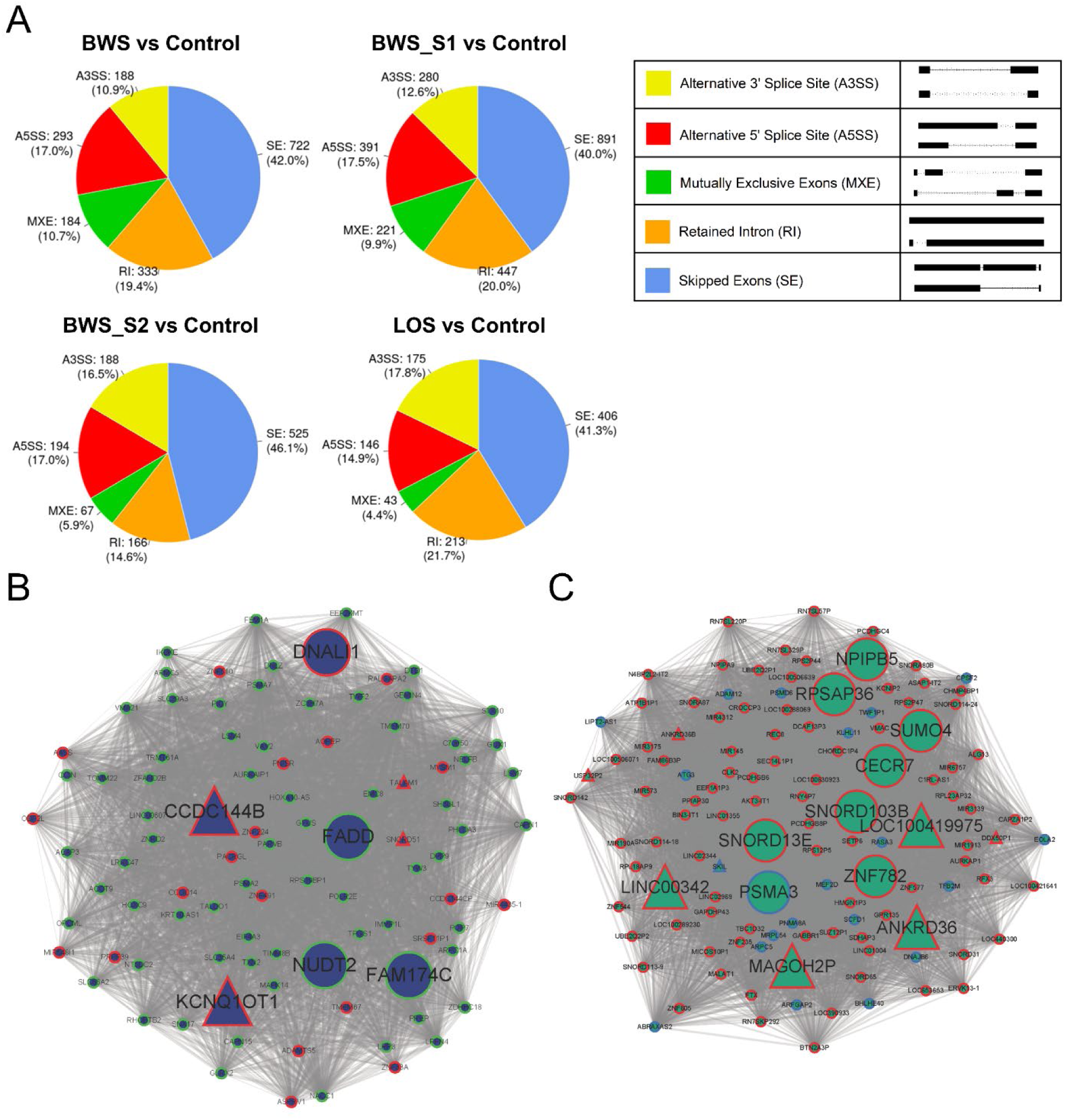
Identification of alternative splicing events in BWS and LOS and correlation network in BWS. **(A)** The types and numbers of alternative splicing events identified in human BWS, BWS subgroup 1, BWS subgroup 2, and bovine LOS when compared to their corresponding control groups. **(B)** and **(C)** Gene interaction network within midnightblue **(B)** and darkturquoise **(C)** modules from weighted gene co-expression network analysis (WGCNA) for BWS subgroup 1. Significantly differentially expressed genes in BWS subgroup 1 are marked with triangles. Red border line of triangles and circles indicate higher gene expression in BWS subgroup 1 regardless of significance, and green/blue border line indicate lower expression. Hub genes are marked with larger triangle/circle.

### Identification of genes associated with clinical features in BWS

To understand the correlation between transcriptional regulation and phenotypic characteristics of human BWS features, weighted correlation network analysis (WGCNA) was performed to determine gene clusters with high correlation in BWS (Figure S4 and Table S2). As a result, for BWS subgroup 1, genes were grouped into 27 modules based on similarity of expression. Several modules showed a significant positive or negative correlation with macroglossia (black, lightcyan, and red) and omphalocele (darkturquoise and midnightblue) suggesting that genes in these modules may serve as biomarkers of these BWS pathologies (Figure S4B). Other suggestive features such as a birthweight greater than two standard deviations (Birthweight_2SDs), facial nevus simplex, and ear creases or pits also showed a significant correlation with some gene modules (Figure S4C). Based on the standard of BWS scoring from Wang et al (65), a comprehensive BWS score was generated for each BWS and control individual and used for WGCNA. Significant positive correlation was observed between BWS score and darkturquoise module, and significant negative correlation was observed in darkgrey, midnightblue, red, and yellow modules (Figure S4A). The signature DEG *KCNQ1OT1* was assigned to the midnightblue module as a hub gene which is defined as a gene with high BWS score correlation and module membership score (absolute value ≥ 0.75 for both; Figure 4B). Other hub genes of this module include *DNALI1*, *CCDC144B*, *FADD*, *NUDT2*, and *FAM174C*. *MYSM1*, which belongs to the midnightblue module, is a transcription coactivator and participates in chromatin remodeling and repression of innate immunity and autoimmunity (66). Genes in midnightblue module showed enriched functions in pathways of neurodegeneration and biosynthesis of amino acids (Table S2). In addition, *MIR145* encodes has-miR-145-5p was enriched in darkturquoise module, which has been reported to be differentially expressed in BWS patients with macroglossia by us (20).

For BWS subgroup 2, genes were grouped into 47 modules. Only the lightsteelblue1 module showed a significant positive correlation with BWS score, and several modules showed a significant positive or negative correlation with cardinal and suggestive features (Figure S4D-F and Table S2). The lightsteelblue1 module has 25 hub genes including *CXCL2*, *CXCL3*, *DNAJB2*, *CORT*, *KCNMB3*, *ETV1*, *GSAP*, *GABBR1*, *MCTS1*, *TMEM258*, and several lncRNA and miRNA genes. *KCNQ1OT1* was assigned to the darkred module without a high BWS score correlation thus not marked as a hub gene.

### Altered transcriptome in LOS exhibits similarities with BWS

For bovine, PCA and hierarchical clustering separates the samples based on treatment (Figure S2E) although not by parental allele (Figure S2F) indicating similar global expression profiles from both alleles. Transcriptome comparison between LOS and controls identified 112 DEGs. As expected, the signature imprinted gene of LOS (*IGF2R*) was found downregulated two-fold in the LOS group. *IGF2R* is normally highly expressed from the maternal allele and its protein is a membrane receptor that mediates the degradation of the paternally expressed fetal growth factor IGF2 (67). Four DEGs were shared between LOS and BWS subgroup 1, namely the upregulated *EMID1*, and the downregulated *SERPINE2*, *LRRC8D*, and *GPX1*.

Allele-specific comparison in the bovine control group identified 122 genes with significant biased expression from one allele, including *IGF2R* which has ∼3X higher expression from maternal allele than paternal allele (Table S2). In addition, *GSTK1*, *MAN1B1*, *GSTZ1*, *ARSG*, *SLC25A29*, *TLDC1*, *PDIA4*, *RWDD2A*, and *NARS* have been reported as imprinted genes or to have biased allelic expression in various mammalian species by other researchers(68–76). *SLC25A29*, together with our previously reported *BEGAIN* (77), show biased paternal expression and are located close to the *DLK1*-*DIO3* imprinted domain, which suggest broader regulatory impacts of this domain in bovine.

Allele-specific DEGs were identified in LOS. *IGF2R* is significantly downregulated (2.4X) from the maternal allele in LOS making it similar in expression level to the paternal allele, and the paternal allele has no change. In addition, among the 122 genes with biased allelic expression in controls, *ECHDC2* has higher paternal expression in controls and is downregulated on the paternal allele in the LOS group, and *IGF1R* has lower maternal expression in controls and is upregulated on the maternal allele in LOS group. *ECHDC2* is involved in branched chain amino acid metabolism and its lower expression increases cell viability (78). *IGF1R* encodes a membrane receptor for the IGF signaling and is positively correlated with growth (79) and observed as highly expressed in Wilms tumor (80).

In bovine, upregulated DEGs in LOS group showed enrichment for both hypo- and hypermethylated DMRs at promoter and hypomethylated DMRs at gene body, and downregulated DEGs showed enrichment for hypermethylated DMRs at promoter and gene body. In addition, 983 AS events were detected in the LOS group when compared to the control group (Figure 4A and Table S2). As in human, skipped exons were the most common AS event. 160 AS events in LOS overlap with BWS subgroup 1 including genes that function in chromatin and cytoskeletal organization such as *TNRC18*, *WAPL*, *HIRA*, *EPB41L2*, and *FNBP4*, and 76 overlap with BWS subgroup 2.

### Characterization of chromosome architecture

Chromosome compartments and topologically associating domains (TADs) are the two fundamental elements of chromosome architecture (24). When considering chromosomes in a linear manner, euchromatin and heterochromatin organize in alternating positions throughout the chromosome. The B compartments are enriched for heterochromatin, mainly localized in the peripheral regions in the nucleus or surrounding the nucleoli, and isolated from areas with enrichment of transcription machineries (such as transcription factories) (81, 82). Oppositely, the A compartments are enriched for euchromatin, mainly localized in the interior of the nucleus, between B compartment regions, and is associated with transcription factories (81, 82). TADs are self-interacting genomic regions within a chromosome and constitute the primary units of interphase chromosome folding. The formation of TADs can either facilitate or block spatial interactions between different chromosomal loci (83) and often involve CTCF and cohesin protein complexes (84).

Hi-C sequencing was performed to determine genome-wide chromosome architecture. We first identified the distribution of chromosome compartment and TADs in human control individuals and found that in the sequenceable autosomal genome there were 13,912 TADs (including sub-TADs) with 46.4% of the genome being A compartment (i.e. positive eigenvalue) and 53.6% being B compartment (negative eigenvalue; Table S3). TAD boundaries show enrichment for predicted CTCF binding sites, public CTCF ChIP-seq peaks, transcription start sites (TSS), CpG islands, and depletion for repetitive sequences (Figure 5B).

**Figure 5.**
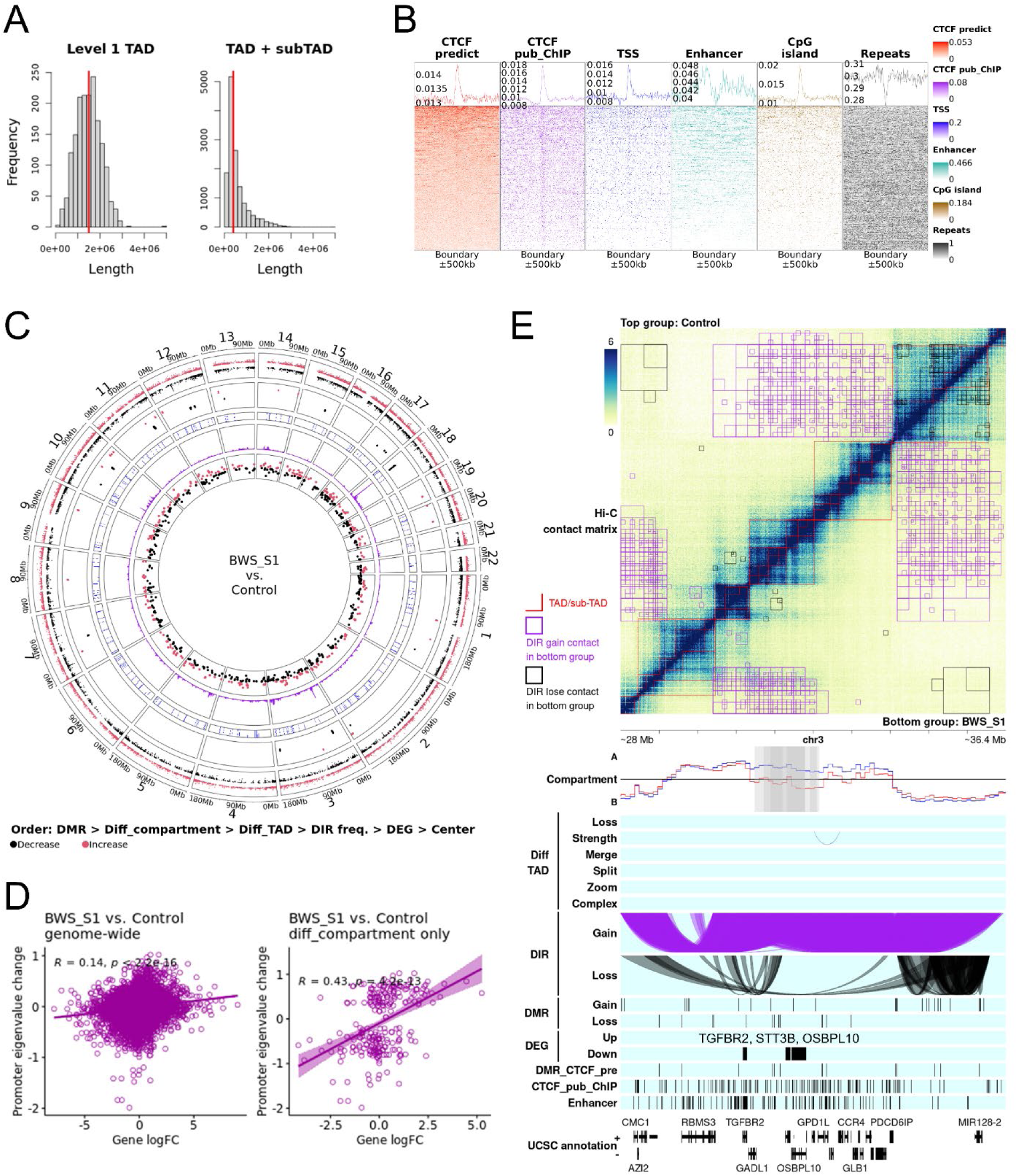
Characterization of human chromosome architecture and alterations in Beckwith-Wiedemann Syndrome (BWS). **(A)** Length distribution of topologically associating domains (TAD) detected in control group. Level 1 TAD refers to the outmost TADs. The red line marks the median length. **(B)** Genomic context enrichment at control group TAD and subTAD boundaries (centered) and flanking regions. Genomic contexts include predicted CTCF binding sites, CTCF public ChIP-seq peaks, transcription start sites (TSS), enhancers, CpG island, and repetitive sequences. The line chart on top of each panel is the average enrichment score for all the boundaries. **(C)** Genome-wide distribution of differentially methylated regions (DMR), differential compartments (diff_compartment), differential TAD (diff_TAD), differential interacting regions (DIR) frequency, and differentially expressed genes (DEG) identified when comparing BWS subgroup 1 with control group. Red dots indicate an increase in CpG methylation, eigenvalue (towards A compartment), and expression level, and black dots indicate decrease. The vertical location of dots indicates the level of changes. Mb = megabases. **(D)** Correlation between chromosome compartment changes (measured by eigenvalue) at gene promoters and log2 transformed fold change (logFC) of protein-coding genes and lncRNA. The left panel shows genome-wide correlations, and the right panel shows correlation at regions of differential compartment. **(E)** Examples of alterations of chromosome architecture associated with gene expression changes in BWS subgroup 1. The top matrix plot shows the Hi-C contacts at 10kb resolution. A deeper blue color indicates higher physical contact frequency. Red lines mark the TADs. All the statistical analyses are conducted by comparing the bottom right group with the top left group. Tracks below the matrix show the location of different elements along this genomic region. The compartment track shows the A/B compartment represented by eigenvalue. The blue and red lines indicate the top left and bottom right group, respectively. The grey area marks significantly differential compartments between groups detected at different resolutions which may overlap and show deeper color. The diff_TAD tracks show the differential TADs in the bottom right group. The DIR tracks show the DIRs that gain or lose contact. The DIRs are also marked in the matrix with purple or black squares. The DMR tracks show DMRs that gain or lose CpG methylation. The DEG tracks show up- or down-regulated DEGs. The DMR_CTCF_pre track shows predicted CTCF binding sites that overlap with DMRs. The CTCF_pub_ChIP track shows the public CTCF ChIP-seq peaks. The enhancer track shows common enhancer regions shared by at least 3 tissues/cell types related to BWS. The UCSC annotation tracks show the UCSC gene annotation.

### Chromosome architecture disruptions in BWS

We detected 14 genomic regions changing towards B compartment (decrease in eigenvalue) and one genomic region changing towards A compartment (increase in eigenvalue) in the BWS group when compared to controls (Figure S5A and Table S4). Only one of these differential compartments (chr17:36000000-36500000) in the BWS group is found in both BWS subgroups, which changes to B compartment with larger eigenvalue decrease in subgroup 1 (-0.7) than 2 (-0.48). This locus is close to but does not overlap with the downregulated gene *DUSP14* in subgroup 1. Other differential compartments in BWS group are found in only one of the two subgroups. For the subgroup-specific analyses, subgroup 1 has 16 and 22 regions changing towards B and A compartments, respectively, and subgroup 2 has 16 and 4 such regions (Figure 5C, S5B, and Table S4). In subgroup 1, 16 of the differential compartments overlap with 29 DEGs and all have the expected direction of changes (e.g. increased eigenvalue and upregulated expression), including *IFIT1*, *IFIT2*, *NAV3*, *ATP2B1*, *FRY*, *PSG5*, *STT3B*, and *SEPTIN7P2*. Permutation tests also verified this significant enrichment (Table S2). The two DEGs in subgroup 2 are not associated with compartment change.

Interestingly, some differential compartments show opposite changes in both subgroups, such as at chr19:42750000-44000000 that has increased and decreased eigenvalue in subgroup 1 and 2, respectively, and overlaps with upregulated *PSG5*, *CEACAM1*, and *PSG10P* in subgroup 1.

For differential TAD analyses, we detected 122, 222, and 49 altered TADs in the BWS group, in subgroup 1, and in subgroup 2, respectively, when compared to controls (Figure 5C, S5A-B, and Table S5). In subgroup 1, 36 DEGs overlap with differential TADs and have no change in chromosome compartments, suggesting altered TADs could contribute to their misregulation. These DEGs include *ATP6V1H*, *FBXL18*, *EPDR1*, *PTP4A1*, *TMEM200A*, *ZMAT3*, *SSR3*, *PEDS1-UBE2V1*, *STK39*, *NOG*, *COPRS*, *SPECC1*, *POMP*, and *OPTN*. For example, altered TADs over *EPDR1* significantly reduce its contact frequency (DIR-Loss track) with neighboring regions which harbor multiple enhancers, which potentially leads to the downregulation of *EPDR1* in BWS subgroup 1 (Figure S7A). In addition, there are 545 differential TADs identified between the two BWS subgroups and overlap with 99 DEGs between them. As illustrated in Figure S7B, *TMEM26* has higher expression in subgroup 1 than 2, and is associated with an altered TAD and higher contact frequency with an enhancer-rich region over *ARID5B* gene body. Overall, in accordance with DNA methylation and transcription results, BWS subgroup 2 possesses a chromosomal structure more similar to that of the control group.

As previously mentioned, both DEGs and DMRs are detected from the *HOX* gene clusters in BWS. The *HOXA* locus is organized by CTCF into chromatin loops and facilitates the stabilization of Polycomb silencing via H3K27me3 (85). All the four *HOX* clusters harbor TAD boundaries (Figure S8). We could not statistically detect the change of CTCF loops within each *HOX* clusters since these loci are too small and we were limited by the resolution of Hi-C sequencing. However, the two neighboring TADs of *HOXA* and *HOXD* showed significant increase in contact frequencies in BWS subgroup 1 when compared to control group, which suggests weakened TAD boundaries within these two clusters (Figure S8). In addition, for *HOXA* region, both BWS subgroup 1 and 2 showed small changes (not significant) of chromosome compartment from control group in opposite direction, and when comparing BWS subgroup 1 to subgroup 2, a significant difference was detected (Figure S9).

### Association between altered chromosome architectures, DEGs, and DMRs in BWS

Pearson correlation tests were performed between eigenvalue changes at gene promoters, which reflect A/B compartment changes, and expression changes of all protein-coding genes and lncRNA (not just DEGs). For genome-wide tests, a very weak positive correlation was observed for BWS subgroup 1 (r = 0.14; Figure 5D). The positive correlation increased in BWS subgroup 1 (r = 0.43) when only considering the differential compartment regions, which indicates that the compartment changes have impacts on gene expression regulation in the differential compartment regions (Figure 5D). BWS subgroup 2 had similar weak correlation at differential compartment regions as its genome-wide correlation (absolute r value < 0.06, figure not shown), indicating the compartment changes have less impact on gene expression in subgroup 2, which is opposite to what was observed for subgroup 1. In addition, BWS subgroup 1 showed a weak positive Pearson correlation between eigenvalue changes and DNA methylation changes at DMRs within the differential compartment regions (r = 0.28; Figure S5D).

These overlapped DMRs are often located in gene bodies, such as the previously mentioned DEG *SEPTIN7P2*, and could reflect changed expression level caused by changes in compartment. However, a solid conclusion cannot be drawn for BWS subgroup 2 since there are only 6 DMRs which overlap differential compartment regions.

Several genomic regions are shown to illustrate the likely impact of altered chromosome architecture on gene expression changes in BWS (Figure 5E and S7). For example, in BWS subgroup 1, *TGFBR2*, *STT3B*, and *OSBPL10* are downregulated and within a differential compartment region that changes toward B compartment (Figure 5E). Further, the downregulated gene *SORBS2* resides in a region that gains contact with surrounding B compartment regions and changes toward B compartment in BWS subgroup 1 (Figure S7C). The promoter of *SORBS2* also overlaps with a hypomethylated DMR (-21.76%; chr4:186877084-186877923). Yet another example, occurs on chromosome 10, in which seven neighboring genes, namely *PAPSS2*, *STAMBPL1*, *FAS*, *LIPA*, *IFIT2*, *IFIT1*, and *PPP1R3C*, are downregulated and associated with a differential change to B compartment (Figure S7D).

As mentioned before, DNA methylation at promoters and enhancers have repressive impacts on gene expression through blocking binding sites for transcription factors (86). The example loci above suggest a model in BWS in which changes of DNA methylation or chromosome compartment will affect gene expression in an independent manner. In the DEG examples illustrated above for the BWS subgroup 1, except *SORBS2*, no DMR was found at the promoters of these DEGs, and their expression changes follow the alterations of their chromosome compartments in which changes towards an A compartment leads to gene upregulation and changes towards the B compartment leads to gene downregulation. In the example of *SORBS2*, its hypomethylated DMR state may further open the promoter for transcription factor binding, however, its spatial change towards B compartment will reduce the availability of transcription machineries in its surrounding microenvironment. So, in this case, DNA methylation and chromosome compartment form an antagonistic situation on gene expression regulation, where the one with a more severe change, or stronger effect, might overcome the effect of the other. This model could explain some of the conflicts between promoter DNA methylation changes and gene expression changes observed in our BWS datasets. In addition, as mentioned above, alterations in TAD and chromosome compartment in BWS could change enhancer availability and contacts with gene promoters and further affect gene expression.

### Conservation of chromosome architectures between human and bovine

The same analyses for contact frequency, chromosome compartment, and TAD were also applied to bovine groups (Figure 6, S6, S10, Table S3, S4, S5, and S6). TAD boundaries in bovine controls show similar enrichment for CTCF, TSS, and CpG islands as in human (Figure 6B). Differential compartments in LOS group also show a positive correlation with gene expression changes as in BWS (Figure 6D). Loci of DEG *KITLG* and *IGF1R* are illustrated as examples for differential compartments and allele-specific architecture in LOS (Figure 6E and S10). We first investigated the conservation of chromosome compartment and TAD between human and bovine genomes. The genomic regions of 1,783 out of 1,940 compartments (91.9%) in human control group can be found in the bovine genome. Over these regions, 882 (49.5%) human compartments were found conserved in bovine control group with greater than 90% overlapping (Figure 7A and Table S7). Similarly, the genomic region of most TADs in human control group can be found in bovine genome (i.e. 96.5%). Over these regions, 15.4% (n = 2,064) of human TADs are conserved in the bovine control group with greater than 90% overlap between species (Figure 7C).

**Figure 6.**
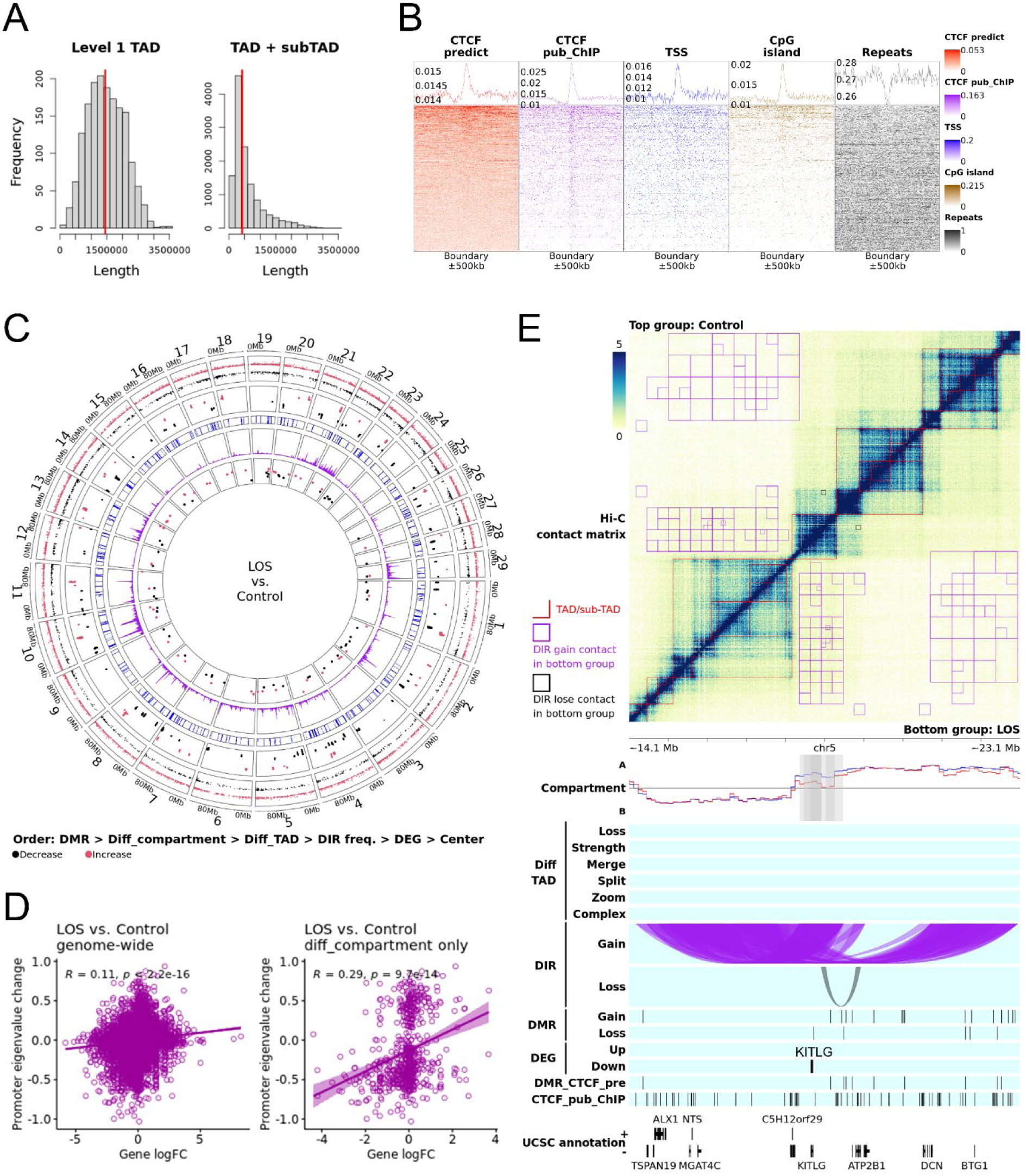
Characterization of bovine chromosome architecture and alterations in large offspring syndrome (LOS). **(A)** Length distribution of topologically associating domains (TAD) detected in control group. **(B)** Genomic context enrichment at control group TAD and subTAD boundaries (centered) and surrounding regions. **(C)** Genome-wide distribution of differentially methylated regions (DMR), differential compartments (diff_compartment), differential TAD (diff_TAD), differential interacting regions (DIR) frequency, and differentially expressed genes (DEG) identified when comparing LOS group with control group. **(D)** Correlation between chromosome compartment changes (measured by eigenvalue) at gene promoters and log2 transformed fold change (logFC) of protein-coding genes and lncRNA. **(E)** Examples of alterations of chromosome architecture associated with gene expression changes in LOS fetuses. Detailed descriptions refer to Figure 5.

**Figure 7.**
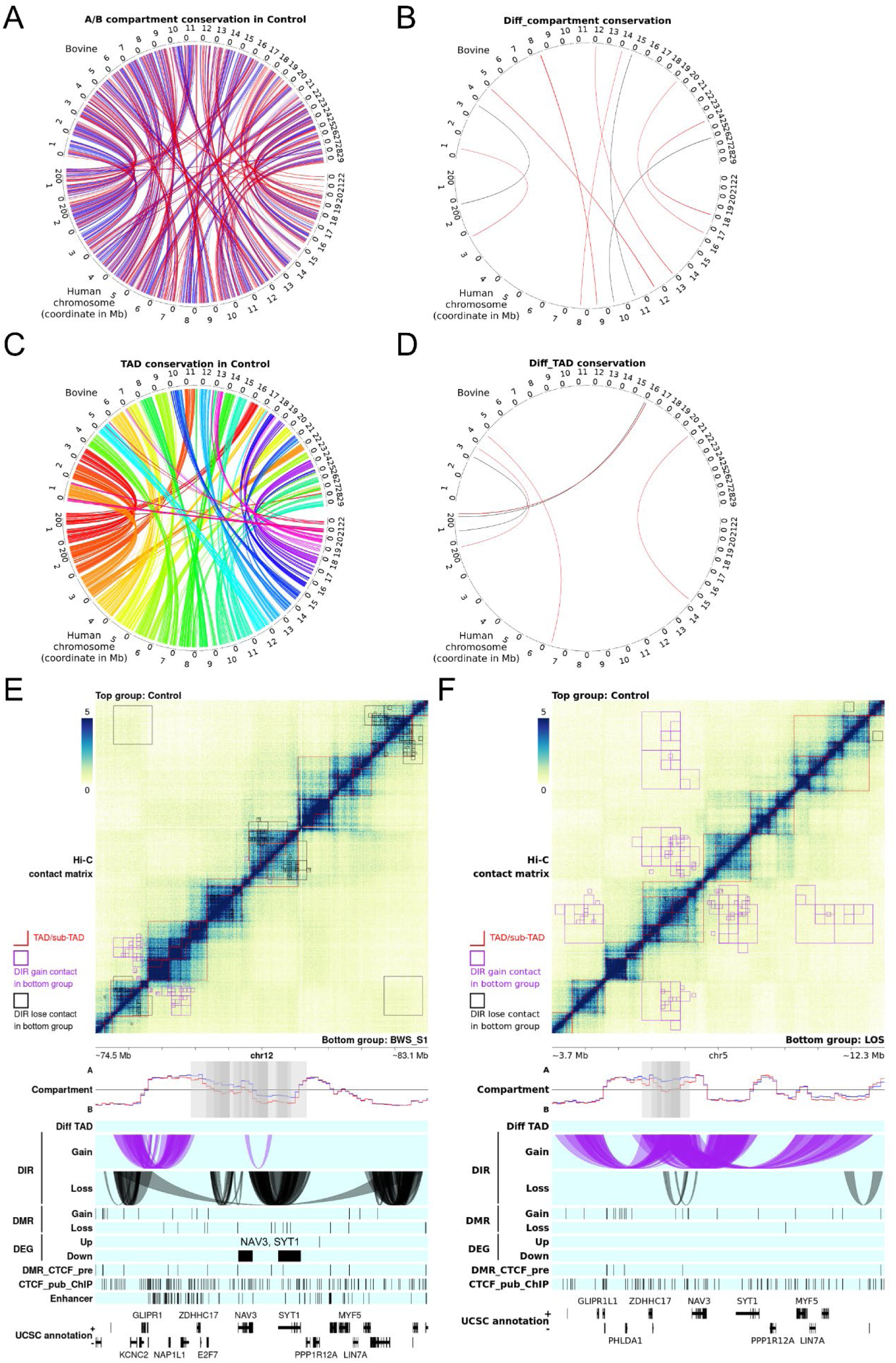
Chromosome architecture conservations between human and bovine. **(A)** Genome-wide conservation of control group A/B compartment between human and bovine. Each line indicates a conserved compartment. Red and blue refer to A and B compartment, respectively. **(B)** Overlapped differential compartments identified in Beckwith-Wiedemann Syndrome (BWS) and large offspring syndrome (LOS). Information regarding comparisons used refer to Methods. Each line indicates an overlapped differential compartment. Red and black refer to differential compartments with same or opposite direction of change between BWS and LOS, respectively. **(C)** Genome-wide conservation of control group topologically associating domains (TAD) between human and bovine. Each line indicates a conserved TAD. Different colors refer to different human chromosomes. Mb = megabases. **(D)** Overlapped differential TAD identified in BWS and LOS. Each line indicates a shared differential TAD between BWS and LOS results with greater than 90% overlap in both species. Green and black refer to shared differential TAD with same and different type between BWS and LOS, respectively. **(E)** and **(F)** An example region of conserved chromosome architecture in control group between human and bovine, and shared compartment changes between BWS and LOS. Detailed descriptions refer to Figure 5E.

Similar approaches were used to find shared differential compartments between BWS and LOS. In total, 16.4% (n = 12) records of differential compartments from human overlap with bovine records (without a cutoff for overlap percentage and regardless of direction of change; Figure 7B and Table S7). Similarly for differential TADs, 2.0% (n = 8) human records overlap with bovine records with a 90% overlap cutoff in both species (Figure 7D). A 9 mb region is shown as an example to illustrate the TAD and compartment conservation and shared compartment changes between human and bovine (Figure 7E and 7F). In control groups, TADs and compartments are largely conserved in this region, and a 1.25 mb differential compartment region that changes toward B compartment is shared by BWS subgroup 1 (chr12:77000000-80000000, half is shared) and LOS (chr5:6000000-7250000) groups. In addition, two downregulated DEGs, namely *NAV3* and SYT1, were identified at this region in BWS. These shared changes of chromosome compartment and TAD between BWS and LOS will be worthwhile for further detailed investigation and have the potential to serve as molecular biomarkers for this syndrome.

## Conclusion

We demonstrate, for the first time, that two molecular subpopulations of BWS IC2 patients exist. We also highlight genes and genomic regions that have the potential to serve as targets for biomarker development to improve current molecular diagnostic methodologies. Our results suggest that genome-wide alternation of chromosome architecture, which is partially caused by altered DNA methylation, also contribute to the development of BWS and LOS. We show shared regions of dysregulation between BWS and LOS, including several *HOX* gene clusters. With the advent of personalized medicine, the bovine model of overgrowth will be invaluable to be able to perform animal and in vitro studies, such as CRISPR/Cas9-based epigenetic editing, of regions identified as altered, in order to develop methodologies to ameliorate the clinical features and treat BWS.

## EXPERIMENTAL MODEL AND SUBJECT DETAILS

### Human patients

Patient skin fibroblast samples and clinical information were collected through the BWS Registry, under the oversight of the Children’s Hospital of Philadelphia (CHOP) Institutional Review Board protocol (IRB 13-010658) and in accordance with the Declaration of Helsinki. In brief, consent was obtained from all patients and/or legal guardians to collect longitudinal clinical information, in addition to samples that became available through clinical care and publish the findings. Patients were identified from the registry cohort and selected based on the availability of skin fibroblasts and their genetic or epigenetic type of BWS. Non-BWS samples (control) were collected for consideration of other growth difference (i.e. have an omphalocele but do not have BWS).

Patient skin fibroblasts were cultured as previously described (29). Briefly, skin samples collected from regions either behind the ear or abdomen were split to process one chemically and other mechanically. Chemical disruption was performed using collagenase and the other was mechanically minced using a scalpel blade. Both explants were seeded into a T25 flask and supplemented with RPMI skin media (RPMI with fetal bovine serum, penicillin–streptomycin antibiotic, and a final concentration of 2 mM L-glutamine). All flasks were incubated at 37°C for up to one month, with periodic media changes. Successful explant cultures were passaged for sustained growth and trypsinized to freeze and stored in liquid nitrogen. Clinical testing for BWS was performed at the University of Pennsylvania Genetic Diagnostic Laboratory as previously described (87).

The identifier and sex of patient included in this study were as follows; 1) control: CON_H_#1 to #3 (female), CON_H_#4 and #5 (male); 2) BWS with IC2 loss of methylation: BWS_#1_IC2 to BWS_#6_IC2 (female), BWS_#7_IC2 to BWS_#12_IC2 (male). The age of fibroblast sample collection (in days with gestational age correction) for each group are control (average 139, range 1-505), BWS with IC2 loss of methylation (327, 1–1283).

### Bovine animals

Day 105 *Bos taurus indicus* (*B. t. indicus*; Brahman breed) x *Bos taurus taurus* (*B. t. taurus*; Angus breed) F1 hybrid fetuses were generated by our laboratory in 2019 and used as tissue donors (30, 88). This breeding strategy introduced genetic variants between the maternal and paternal alleles for allele-specific analyses. The control group was generated using artificial insemination and the test ART group was generated by *in vitro* production procedures. The LOS group was defined as individuals from the ART group with body weight greater than 97^th^ centile of controls (548g). Conceptuses were collected by caesarean section to maintain nucleic acid integrity. Fetal skin fibroblast primary cell lines were established and maintained as described in our previous study (30). The identifier, original ID, sex, and body weight of fetuses used in this study were as follows; 1) control fetuses: CON_B_#1 (original ID 533, female, 388g), CON_B_#2 (647, female, 396g), CON_B_#3 (640, male, 448g), CON_B_#4 (648, male, 466g), and CON_B_#5 (527, male, 514g); 2) LOS fetuses: LOS_#1 (616, female, 638g), LOS_#2 (656, female, 704g), LOS_#3 (604B, female, 986g), LOS_#4 (512, male, 648g), LOS_#5 (602, male, 752g), and LOS_#6 (664, male, 1080g). It should be noted that one of the LOS samples, LOS_#1, is considered an outlier according to all the sequencing results, therefore it is excluded from the results presented in the main document (LOS group). Statistical comparisons including LOS_#1 are labeled as LOS_All.

All the animal procedures were approved by the University of Missouri Animal Care and Use Committee under protocol 9455. Trained personnel and Veterinarians performed all animal handling and surgeries.

## METHOD DETAILS

In this manuscript, all the chromosomal coordinates for bovine genome refers to assembly ARS-UCD1.2 (bosTau9), and for human genome refers to GRCh37 (hg19) (89).

### Genomic DNA extraction

Genomic DNA from human skin fibroblasts was isolated using the AllPrep DNA/RNA Micro Kit (QIAGEN) per the manufacturer’s instructions, as previously described (29). Briefly, the cells were lysed in the Buffer RLT Plus. The lysate was then homogenized into a QIAshredder spin column. The homogenized lysate was then transferred to an AllPrep DNA spin column, washed with washing buffer and then DNA was eluted using elution buffer. The concentration of DNA was measured by using a NanoDrop® ND-1000 Spectrophotometer (Thermo Fisher Scientific). Genomic DNA samples were stored at -20°C.

Bovine fibroblast cells were lysed in lysis buffer (0.05 M Tris-HCl (pH 8.0), 0.1 M EDTA, and 0.5% (w/v) SDS) with proteinase K (Fisher BioReagents, BP1700) at 55°C for four hours. Genomic DNA was extracted with Phenol:Chloroform:Isoamyl Alcohol (SIGMA, P3803) following the manufacturer’s instructions. The concentration of DNA was measured by using a NanoDrop® ND-1000 Spectrophotometer and DNA integrity was confirmed by electrophoresis on a 0.7% agarose gel. Genomic DNA samples were stored at -20°C.

### RNA isolation

Total RNA from human skin fibroblasts was isolated using the AllPrep DNA/RNA Micro Kit (QIAGEN) per the manufacturer’s instructions, as previously described (29).

Briefly, the cells were lysed in the Buffer RLT Plus. The lysate was then homogenized into a QIAshredder spin column. The homogenized lysate was then transferred to an AllPrep DNA spin column, and the flowthrough was used for RNA extraction using 70% ethanol. The concentration of RNA was measured by using a NanoDrop® ND-1000 Spectrophotometer and RNA samples were stored at -80°C.

Total RNA was isolated from bovine fibroblast cells using TRIzol™ Reagent (Invitrogen, 15596026) following the manufacturer’s instructions. The concentration of RNA was measured by using a NanoDrop® ND-1000 Spectrophotometer. RNA samples were stored at -80°C.

### Hi-C sequencing and data analyses

Hi-C sequencing for fibroblast cells was conducted by CD Genomics. Information on library preparation and sequencing obtained from the company is as follows: Cells were cross-linked with 1% formaldehyde at room temperature and quenched with glycine. The cross-linked cells were subsequently lysed. Endogenous nucleases were inactivated with 0.3% SDS, then chromatin DNA were digested by MboI (NEB), marked with biotin-14-dCTP (Invitrogen), and ligated by T4 DNA ligase (NEB).

After reversing cross-links, the ligated DNA was extracted through QIAamp DNA Mini Kit (Qiagen) according to manufacturers’ instructions. Purified DNA was sheared to 300-500 bp fragments and were further blunt-end repaired, A-tailed and adaptor-added, followed by purification and PCR amplification. Finally, equimolar pooling of libraries was performed based on QC values and sequenced on Novaseq6000 sequencer (Illumina) with a read length configuration of 150 PE for 1,000M PE reads per sample (1,000M in each direction).

The processing of Hi-C raw sequencing reads was based on the ‘Hi-C Processing Pipeline’ from 4D Nucleome Data Portal (https://data.4dnucleome.org/resources/data-analysis/hi_c-processing-pipeline). Briefly, reads were aligned to the corresponding reference genome using bwa 0.7.17 with parameters ‘mem -SP5M’ (90). Reads with mapping quality (MAPQ) equal or greater than 30 were kept for analyses. Then, pairtools 0.3.0 was used to parse, sort, and deduplicate aligned reads, and pair types ‘UU’, ‘UR’, and ‘RU’ were selected (91).

Cooler and Juicer were used to convert contact matrix to .cool and .hic files (92, 93). Hi-C data was also used to identify genomic variants following the pipeline for 1000 bull genome project (94). Briefly, Read groups were added using AddOrReplaceReadGroups function of picard 2.25.5 (95) for aligned Hi-C sequencing reads. Duplicated reads were marked using MarkDuplicates function of picard with parameter ‘--OPTICAL_DUPLICATE_PIXEL_DISTANCE 2500’. For human, the dataset of known variants was acquired from GATK resource bundle (https://gatk.broadinstitute.org/hc/en-us/articles/360035890811-Resource-bundle). For bovine, the dataset of known variants was acquired from the 1000 bull genome project, namely ARS1.2PlusY_BQSR_v3.vcf.gz. GATK 4.2.6.1 (96) was used to recalibrate base quality and identify genomic variants in the Hi-C data with the known variant dataset as reference. Parameters used for BaseRecalibrator were ‘–bqsr-baq-gap-open-penalty 45 --read-filter MappingQualityReadFilter --minimum-mapping-quality 20’ for bovine. For human, the same settings were used except with default value for --bqsr-baq-gap-open-penalty. In addition, for bovine, our previously generated genomic sequencing data for semen DNA of the bull used to sire all the fetuses in this study was included for joint genotyping in order to assign alleles for the genomic variants (GEO: GSE197130) (30). HaplotypeCaller was used with parameter ‘-ERC GVCF’, and GenomicsDBImport and GenotypeGVCFs were used with default settings. For bovine, raw SNP and INDEL were separately scored using VariantRecalibrator with parameter ‘--resource:1000G,known=false,training=true,truth=true,prior=10.0 known_variant -an QD -an MQ -an MQRankSum -an ReadPosRankSum -an FS -an SOR’ was used. For human, parameter ‘--resource:hapmap,known=false,training=true,truth=true,prior=15.0 known_variant1 --resource:omni,known=false,training=true,truth=true,prior=12.0 known_variant2 --resource:1000GSNP,known=false,training=true,truth=true,prior=10.0 known_variant3 --resource:1000GINDEL,known=false,training=true,truth=true,prior=10.0 known_variant4 --resource:dbsnp,known=true,training=false,truth=false,prior=2.0 known_variant5 -an QD -an MQ -an MQRankSum -an ReadPosRankSum -an FS -an SOR’ was used. Scored variants were filtered using ApplyVQSR with parameter ‘--truth-sensitivity-filter-level 99.0’.

For bovine, variants used for allele assignment met the following criteria: 1) marked as PASS by ApplyVQSR, 2) GQ>=40 in the bull and the fetus, 3) DP>=10 in the bull, 4) no read supports not assigned alleles in the bull and the fetus, 5) only one allele of the bull is shown in the fetus. In addition, variants on chrX identified in male fetuses were excluded from analyses, since which is likely due to the lack of chrY in the reference genome. Alleles were assigned for bovine Hi-C reads based on qualified variants using custom Perl scripts. For SNP, the nucleotide in the reads at the SNP location was directly used for allele assignment. For INDEL, local maternal and paternal allele pseudo genomes were generated, and the reads were realigned to both pseudo genomes. The reads were assigned to the alleles with more aligned bases, or less mismatches if aligned base numbers were the same. Next, variants with biased allelic ratio of Hi-C reads (one allele is less than 15%) were excluded from analyses. Lastly, reads that cover multiple variants and have conflicting allele assignments were excluded from the analyses.

Statistical analyses for detecting differentially interacting regions (DIR), differential TAD, and differential chromosome compartments were performed. The sex chromosomes were excluded from analyses to circumvent confounding created by X chromosome inactivation. R package multiHiCcompare 1.12.0 was used to identify DIR at 1mb, 500kb, 250kb, 100kb, and 50kb resolutions between groups (97). The make_hicexp function was used with default parameters except ‘remove.regions = NULL’, and cyclic_loess and hic_exactTest functions were used with default settings. False discovery rate (FDR) was controlled at 0.05 for significance.

For visualization and TAD detection for each group, the size of contact matrix was normalized to the first sample in the Control group for both human (CON_H_#1) and bovine (CON_B_#1), and group mean contact matrix was calculated based on normalized matrix of each sample in the group. R package SpectralTAD 1.10.0 was used with parameter ‘qual_filter = TRUE, z_clust = FALSE, levels = level, resolution = 25000, window_size = window_size, gap_thresh = 0.2, min_size = 6’ to identify TAD structure ranging from 150kb to 4mb at 25kb resolution (98). Level and window_size was initially set to 10 and 120, respectively, and reduced by 1 and 30 each time if error reported. In addition, diffDomain was used with function and parameter ‘dvsd multiple --min_nbin 1 --hicnorm NONE’ and ‘adjustment fdr_bh’ to detect differential TAD boundaries at 25kb resolution between groups based on TAD identified by SpectralTAD (99). Since SpectralTAD and diffDomain only support single sample comparison instead of group comparison, to reduce the impacts of sample variations within each group and obtain more consistent differences between groups, filtering for inconsistent TADs of both groups involved in comparison was performed for detected differential TADs. Inconsistent TADs were obtained by applying diffDomain between each sample and its group mean contact matrix and merging all the differential results from each sample. Types of differential TAD include loss, strength change, merge, split, zoom, and complex. Strength change refers to the situation that the same TAD is detected in the two groups for comparing but the chromosomal contact frequency is different within the TAD. TAD with strength change could reflect forming or losing small sub-TADs and loops within it that requires higher resolution and cannot be properly annotated with the sequencing depth in this study. TAD with strength change could also be a result of chromosome compartment change of the sub-TADs within it. Zoom refers to the extension or shrink of a TAD in another group. Complex refers to the situation that cannot be simply categorized into any other type.

dcHiC was used to assign chromosome compartments and identify differential compartments at 1mb, 500kb, 250kb, and 100kb resolutions between groups (100). Default settings for cis, select, analyze, and subcomp functions of dcHiC were used, and FDR was controlled at 0.05 for significance during the analyze step. Principle component (PC) selection for some chromosomes was assigned manually instead of using the results of the select function. This was because for certain chromosomes, the score (correlation with CpG and gene) of PC1 and PC2 are very similar, and auto PC selection assign PC1 for one group and PC2 for another group, which results in large amount of false positive differential compartments. For a given chromosome, manual PC selection was performed as follow. First, for each sample, if the difference between PC1 and PC2 scores was less than 30% of the highest PC score (normally it’s larger than 50%), the sample was considered having similar PC score. Second, for samples with similar PC score, if more than half samples were also considered having similar PC score, the PC with the highest score from the majority of samples would be applied, otherwise the auto selected PC would be applied. Last, for samples without similar PC score, always the PC with the highest score was selected. In addition, for differential compartment results, a filter was applied to further remove false positive results associated with unmappable regions. The filter was set as that for any significant differential compartment, if it had no overlap with any significant results from the other three resolutions, it would be removed. The quantile normalized principal component (eigenvalue) calculated at 100kb resolution was used for assigning compartments and plotting. In each group, continuous 100 kb bins with the same A or B compartment assignment were merged to generate the final compartment list.

### Whole genome bisulfite sequencing (WGBS) and data analyses

WGBS for fibroblast cells was conducted by CD Genomics. Information on library preparation and sequencing obtained from the company is as follows: Genomic DNA was fragmented by sonication to a mean size of approximately 200-400 bp.

Fragmented DNA was end-repaired, 5’-phosphorylated, 3’-dA-tailed and then ligated to methylated adapters. The methylated adapter-ligated DNAs were purified using 0.8× Agencourt AMPure XP magnetic beads and subjected to bisulfite conversion by ZYMO EZ DNA Methylation-Gold Kit (Zymo Research). The converted DNAs then have the addition of 8-bp index primers to create the final cDNA library. Equimolar pooling of libraries was performed based on QC values and sequenced on Novaseq6000 sequencer (Illumina) with a read length configuration of 150 PE for 300M PE reads per sample (300M in each direction). 0.1-1% lambda DNA were added during the library preparation to monitor bisulfite conversion rate.

For WGBS data, raw sequencing reads were deduplicated using the clumpify function of BBMap 38.90 (101) and trimmed for adapter sequences and low quality bases using trimmomatic 0.39 (102) with parameters ‘ILLUMINACLIP:adapter_seq:2:30:10:1:true LEADING:20 TRAILING:20 AVGQUAL:20 MAXINFO:0:0.5’. Trimmed reads were aligned to the corresponding reference genome using BSBolt 1.5.0 (103) with default setting. Trimmed reads were also aligned to lambda phage genome to determine bisulfite conversion rates. Samtools 1.13 (104) was used to convert, sort, filter, and index bam files. MarkDuplicates function of picard 2.25.5 (95) was used with parameter ‘--OPTICAL_DUPLICATE_PIXEL_DISTANCE 2500’ to further remove duplicated reads after alignment. CpG methylation information was extracted from the bam files using the function CallMethylation of BSBolt with parameter ‘-CG -min 1 -BQ 20’. For both human and bovine samples, CpG methylation at C/T SNPs detected in their corresponding Hi-C data were excluded for sense strand reads, and CpG methylation at G/A SNPs were excluded for antisense strand reads. For bovine samples, SNPs identified from Hi-C dataset were also used to assign alleles for WGBS reads using custom Perl scripts. C/T SNPs (based on sense strand sequence) were only applied for antisense strand reads, and G/A SNPs were only applied for sense strand reads. Other types of SNP were applied for both strands. Reads that cover multiple SNPs and have conflicting allele assignments were excluded. For both overall and allele-specific comparisons, statistical analyses were conducted using R package hummingbird (105) with parameter ‘minCpGs = 10, minLength = 100, maxGap = 300’ to identify differentially methylated regions (DMRs) between groups. DMRs with at least 15% difference in methylation level (both gain and loss of methylation) and at least 4 mean read coverage at CpG sites in all the samples were reported. For allele-specific analyses, the read coverage filtering was not applied since only about 20% of the usable SNPs are shared by all the sample. The sex chromosomes were excluded from analyses to circumvent confounding created by X chromosome inactivation.

### Total RNA sequencing (total RNA-seq) and data analyses

Total RNA-seq for fibroblast cells was conducted by CD Genomics. Information on library preparation and sequencing obtained from the company is as follows: The first step involves the removal of ribosomal RNA using RiboZero kit (Illumina).

Subsequently, the RNA is fragmented into small pieces using divalent cations under elevated temperatures. The cleaved RNA fragments are copied into first strand cDNA using reverse transcriptase and random primers, followed by second strand cDNA synthesis. These cDNA fragments then have the addition of a single ’A’ base and subsequent ligation of the adapter. The products are purified and enriched with PCR to create the final cDNA library. This results in libraries with inserts ranging in size from 120–200 bp with a median size of 150 bp. Equimolar pooling of libraries was performed based on QC values and sequenced on Novaseq6000 sequencer (Illumina) with a read length configuration of 150 PE for 33M PE reads per sample (33M in each direction).

All read data was assessed for quality with FastQC 0.11.7 and low-quality bases (Phred scores ≤ 20) were trimmed using the dynamictrim function of SolexaQA++ 3.1.7.1. Reads less than 60 bases in length were removed using SolexaQA++ LengthSort. Known splice sites were extracted from the gtf file of each reference genome using the Python script hisat2_extract_splice_sites.py. Hisat2 2.1.0 was used to establish a genome index with the hisat2-build and also to align the quality trimmed reads to the reference genome with the following adjusted parameters: --mp 6,6; – score-min L,0,-0.2; known-splicesite-infile. Sam files were converted to bam files and coordinate-sorted using Samtools 1.8. Then, Stringtie 2.1.4 was used with the sorted bam files to assemble transcriptomes for each sample. Stringtie merge was used to combine the assembled transcripts from all samples into a reference assembly. Assembled transcripts were extracted using the gffread module in cufflinks 2.2.1. An index was built for the merged transcriptome and reads were quantified using Salmon 0.11.1. IsoformSwitchAnalyeR 1.17.04 was used to rescue stringtie annotations and generate a gene count matrix.

For alternative splicing (AS) event analyses, the merged Stringtie annotation file and coordinate-sorted bam files were used, and AS events were called using rMATs 4.3.0 with parameters ‘-t paired –readlength 150 -cstat 0.0001 -libType fr-firststrand --allow-clipping --variable-read-length’. Results of JCEC version were used. FDR was controlled at 0.05 for significance.

To assign allele specificity for total RNA-seq reads in bovine, variants were called from the aligned data according to GATK’s Best Practices for Calling Variants in RNA-seq data. Duplicate reads were removed and read groups were assigned using Picard 2.22.4. A dictionary was created for the reference genome using GATK’s CreateSequenceDictionary. GATK SplitNCigarReads was used to hard clip sequences extending into intronic regions and GATK BaseRecalibrator to recalibrate base quality scores. Then, HaplotypeCaller was run on calibrated data and called variants were filtered with adjusted parameters (AB < 0.2 & MQ0 > 50). Variants that were shared between genomic and RNA-seq data for each sample were kept. Variants that were heterozygous in the fetus and homozygous in the bull were used to assign alleles. A variant annotation file was created using the annotate function of bcftools 1.8 and gene annotations were added using custom scripts. Custom R scripts were used to retrieve maternal and paternal allele depth read counts from VCF files in order to quantify allele-specific expressions. Genes with reads covering at least one informative SNP (needed to assign alleles) in all samples were included for analyses. The raw allelic read counts for each gene in each sample were further used to calculate allelic ratios and multiplied by the total (non-allelic) gene read counts. This step was included since only ∼20% of the informative SNPs were shared by all samples and the raw allelic read counts for genes would be largely affected by the total informative SNP number in each sample. The allelic ratio adjusted gene read counts were used in downstream statistical analyses.

Statistical analyses were conducted using R package edgeR 3.36.0 (106) to identify differentially expressed genes (DEGs) between groups. Parameters used include ‘method = “TMM”’ for calcNormFactors function and ‘robust = TRUE’ for glmFit function. FDR was controlled at 0.05 for significance.

### Functional correlation between gene expression and BWS clinical diagnosis

To determine the co-expressional network that gathered by genes showing similar trends across samples, WGCNA was performed in R (107). Briefly, gene abundance matrix (normalized expression matrix) from IsoformSwitchAnalyeR was log2 transformed and only genes with high expressional variability values (top 60%, 15304 genes retained) were remained for identification of co-expression network and a soft threshold β value was set as 10 according to the criterion of approximate scale-free topology. BWS specific clinical diagnosis was generated through the evaluated features from Wang et al reported study (65). Clinical BWS score consists of the combination of cardinal and suggestive features, with one more point (plus two points) was given for each diagnosed cardinal feature, while one point was given for each diagnosed suggestive feature. The correlation between eigengene (a representative of the gene expression profiles in a module) in each module and clinical diagnoses was calculated using Pearson’s correlation method, and only relationships with p-value < 0.05 were considered as significantly positive/negative correlation. Hub genes within BWS-related module were identified with both absolute gene-BWS score significance ≥ 0.75 and absolute module membership ≥ 0.75.

### Correlation analyses among DNA methylation, gene expression, and chromosome compartment

Pearson correlation analyses for comparison 1) between DNA methylation changes and eigenvalue changes and 2) between gene expression changes and eigenvalue changes at gene promoter were performed using the R base function ‘cor’. For gene expression correlation, it should be noted that small RNA genes were not included in the test due to the uncertainty of their promoter locations.

### Genomic content overlap and enrichment analyses

Gene annotation information was obtained from NCBI (GCF_002263795.1_ARS-UCD1.2_genomic.gtf and GCF_000001405.25_GRCh37.p13_genomic.gtf) (108). Repeated and overlapped exons were merged for each gene, and introns were calculated based on merged exons. Promoters (1kb) were calculated based on transcription start sites annotation and only included protein coding genes and long non-coding RNAs. Annotation of CpG islands and repeated sequences were obtained from UCSC Genome Browser (109). Locations of CpG shores (flanking 2kb from CpG islands) and shelves (flanking 2-4kb from the CpG island) were calculated based on CpG island annotation.

Potential CTCF binding sites were predicted globally for human and bovine genome using TFBSTools 1.26.0 (110) with database JASPAR2020 (111). CTCF motifs of ‘vertebrates’ were used for prediction and min.score was set to 81%. Human public CTCF ChIP-seq data of fibroblast primary cells from different tissues were obtained from the ENCODE portal with the following identifiers: ENCFF148VQH (lung), ENCFF337WIE (foreskin), ENCFF344UBE (cardiac), ENCFF438XHB (mammary gland), ENCFF570FLB (lung), ENCFF638OUB (dermis), ENCFF652MLT (foreskin), ENCFF734DZF (lung), ENCFF738CXX (villous mesenchyme), ENCFF859PRV (aortic adventitia), and ENCFF933VBD (pulmonary artery) (112). Peaks from these files were merged to use as common CTCF sites. Bovine public CTCF ChIP-seq data of different tissues (no fibroblast primary cell data available) were obtained from the Gene Expression Omnibus (GEO) database with the following identifiers: GSE158430 (replicate for adipose, cerebellum, cortex, hypothalamus, liver, lung, muscle, and spleen) and GSE129423 (control rumen epithelial primary cells) (113). GSE129423 was based on genome build UMD3.1.1 and was converted to ARS-UCD1.2 using liftOver from UCSC Genome Browser with parameter ‘-minMatch=0.1’. Due to lower quality (i.e. occasionally very broad peaks) of these data than the human data, peak regions found in at least four files were kept as common CTCF sites.

The genomic coordinates of human enhancers were obtained from the EnhancerAtlas2 database (114). 18 tissues/cell types related to BWS from the database were used, which are Fetal_heart, Fetal_placenta, Hepatocyte, Left_ventricle, Myotube, Skeletal_muscle, Fetal_kidney, Fibroblast_foreskin, Kidney, Liver, Pancreas, Spleen, Fetal_muscle_leg, Heart, Kidney_cortex, Mesendoderm, Pancreatic_islet, and Trophoblast. Common enhancer regions were selected with the similar approach used for public CTCF ChIP peaks that regions found in at least three files were kept and resulted in 73473 common enhancers. The genomic coordinates of ICRs in human and bovine referred Jima and Wyss studies (115, 116).

Bedtools and custom Perl scripts were used to identify overlapped genomic contexts for regions with altered DNA methylation, gene expression, and chromosome architecture, and calculate observed frequencies (Obs) (117). Permutation tests were performed to determine expected frequencies (Exp) of overlapping by randomly shuffling genomic contexts across the genome for 10,000 times. For each test, the p value was calculated as p = n(|Exp - mean(Exp)| ≥ |Obs - mean(Exp)|)/10,000. EnrichedHeatmap was also used for enrichment visualization in the heatmaps and different flanking distance was chosen based on the length of genomic content examined to best show the background level and enrichment pattern (118).

### Conservation analyses

For TAD/compartment conservation analyses between human and bovine control groups, the genomic coordinates of human TAD/compartment were converted to bovine coordinates using liftOver from UCSC Genome Browser with parameter ‘- minMatch=0.1’. Then, custom Perl scripts were used to identify bovine TAD/compartment overlapping with human TAD/compartment that the overlapped region is greater than 90% in both species for TAD and in any species for compartment.

To identify shared differential compartments between BWS and LOS, for each comparison, bedtools was first used to merge significant results of different resolutions into non-overlapping regions (117). Merged results of comparisons that compare BWS/LOS to control group were combined for each species. For human, BWS vs Control (index 1 in Table 1), BWS_S1 vs Control (2), BWS_S2 vs Control (3) were used for analyses. For bovine, LOS vs Control (5) was used for analyses. Then human coordinates were converted to bovine using liftOver and regions overlapped with bovine differential compartments were identified using custom Perl scripts.

**Table 1.** Summary of statistical comparisons.

|  |  |  |  |  |  |  |  | Diff_compartment |  |
| --- | --- | --- | --- | --- | --- | --- | --- | --- | --- |
| Species | Type | Index | Comparison | DMR | DEG | DIR* | Diff_TAD | Raw* | Unique |
| Human | Overall | 1 | BWS vs Control | 7865 | 8 | 1399 | 122 | 124 | 15 |
|  |  | 2 | BWS_S1 vs Control | 23582 | 742 | 54921 | 222 | 380 | 38 |
|  |  | 3 | BWS_S2 vs Control | 1915 | 2 | 116 | 49 | 110 | 20 |
|  |  | 4 | BWS_S1 vs BWS_S2 | 20657 | 484 | 68396 | 545 | 548 | 55 |
| Bovine | Overall | 5 | LOS vs Control | 10860 | 112 | 20200 | 299 | 1227 | 136 |
|  |  | 6 | LOS_All vs Control | 4071 | 6 | 566 | 161 | 679 | 82 |
|  | Allele specific | 7 | LOS_Maternal vs Control_Maternal | 17611 | 29 | 1000 | 667 | 342 | 51 |
|  |  | 8 | LOS_Paternal vs Control_Paternal | 15708 | 16 | 1115 | 636 | 309 | 45 |
|  |  | 9 | Control_Paternal vs Control_Maternal | 10143 | 122 | 4940 | 105 | 240 | 31 |
|  |  | 10 | LOS_All_Maternal vs Control_Maternal | 15405 | 2 | 275 | 690 | 140 | 23 |
|  |  | 11 | LOS_All_Paternal vs Control_Paternal | 14061 | 0 | 291 | 675 | 93 | 19 |
| DMR = differentially methylated regions; DEG = differentially expressed genes; DIR = differential interacting regions; diff_TAD = differential topologically associating domains; Diff_compartment = differential compartments; Unique = after merging overlapped and continuous regions with the same direction of change; *Count includes results from all the bin sizes |  |  |  |  |  |  |  |  |  |

### Plot generation

R package ComplexHeatmap, EnrichedHeatmap, plotgardener, circlize, methylKit, ggpubr, ggfortify, ggplot2, VennDiagram, and Python library matplotlib were used for making plots (118–127).

## Data availability

All the raw sequencing data for human samples, including whole genome bisulfite sequencing, total RNA sequencing, and Hi-C sequencing, are available in the database of Genotypes and Phenotypes (dbGaP) with accession number (phs001794.v1.p1). The subject ID of each sample used in dbGaP can be found in Table S8. Some of the patients in this study were also included in our previous study and deposited under the same dbGaP subject ID (20). All the raw sequencing data for bovine samples, including whole genome bisulfite sequencing, total RNA sequencing, and Hi-C sequencing, are available in the Gene Expression Omnibus (GEO) database with accession number (GSE251961).

## Code availability

All original codes have been deposited at Zenodo (https://doi.org/10.5281/zenodo.15060272) and is publicly available as of the date of publication.

## Acknowledgements

We acknowledge the ENCODE Consortium and the John Stamatoyannopoulos (UW) and Bradley Bernstein (Broad) laboratories that generated the human CTCF ChIP-seq datasets. This research was partially funded by the United States Department of Agriculture-Agriculture and Food Research Initiative (USDA-AFRI) grant 2018-67015-27598 (R.M.R. and D.E.H.), by the National Science Foundation grant CCF 2343612 (J.C.), by the National Cancer Institute grant K08 CA193915 (J.M.K.), by Alex’s Lemonade Stand Foundation (J.M.K.), by a Damon Runyon Clinical Investigator Award provided by the Damon Runyon Cancer Research Foundation (105–19; J.M.K.), by the Lorenzo “Turtle” Sartini Jr. Endowed Chair in Beckwith-Wiedemann Syndrome Research (J.M.K.), and by the Victoria Fertitta Fund through the Lorenzo “Turtle” Sartini Jr. Endowed Chair in Beckwith-Wiedemann Syndrome Research (J.M.K.). The funders had no role in study design, data collection and analysis, decision to publish, or preparation of the manuscript.

## Author contributions

Y.L., J.M.K., D.E.H., J.C., and R.M.R. designed research; Y.L. and S.N. performed research; Y.L., P.X., F.B., and A.K.G. analyzed data; Y.L., P.X., F.B., A.K.G., S.N., and R.M.R. wrote the paper; all authors edited the paper.

## Declaration of interests

The authors declare no competing interests.

## Authors’ ORCID

Yahan Li

Ping Xiao

Frimpong Boadu

Anna Goldkamp

Snehal Nirgude

Jianlin Cheng

Darren Hagen

Jennifer Kalish

Rocío Melissa Rivera

---

**Figure S1.**
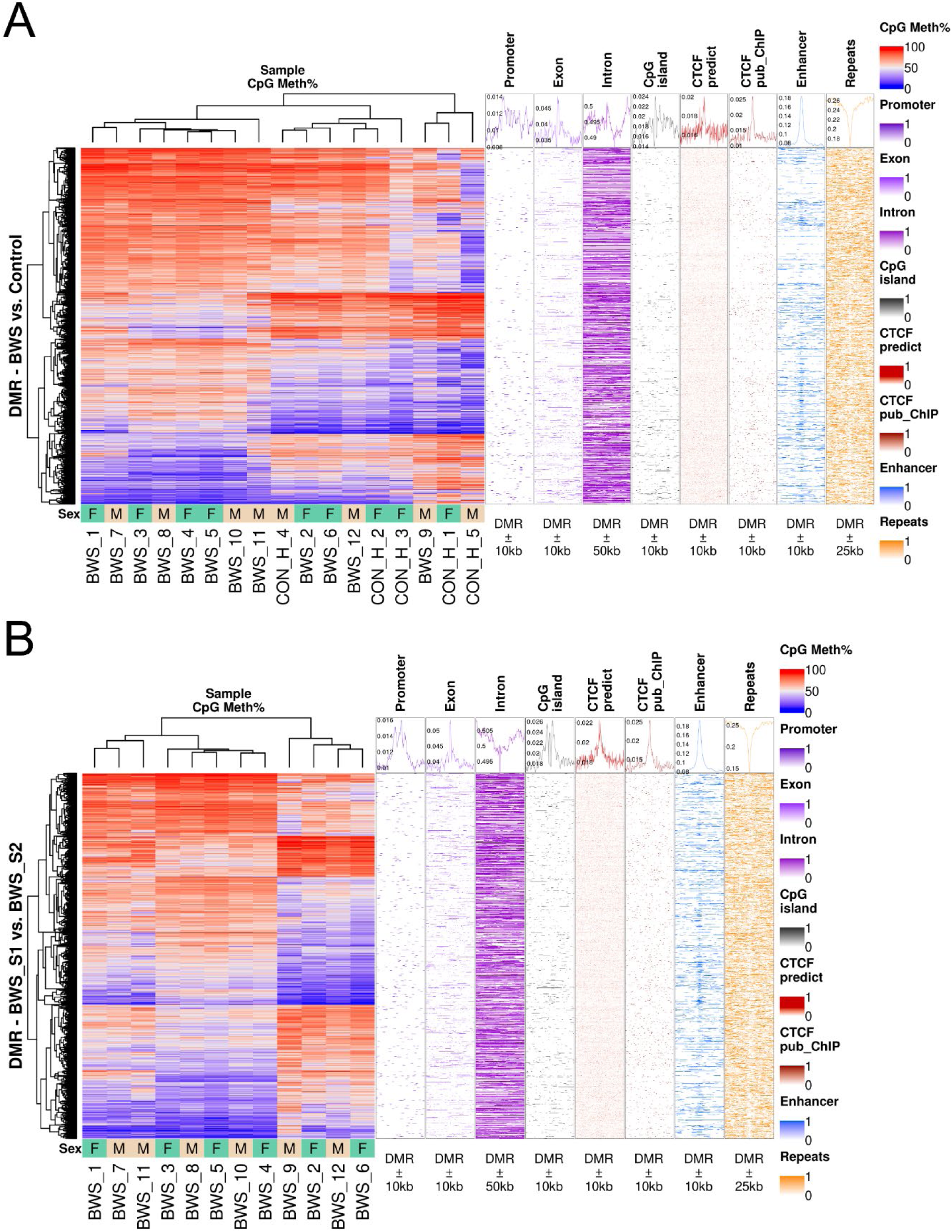
CpG methylation heatmap and genomic context enrichment for differentially methylated regions (DMR) in humans. DMR for human BWS group compared with control group **(A)** and BWS subgroup 1 group compared with BWS subgroup 2 **(B)**. BWS subgroup 1 include BWS #1, #3, #4, #5, #7, #8, #10, and #11, and BWS subgroup 2 include BWS #2, #6, #9, and #12. The left panel is the heatmap with hierarchical clustering and sex annotation which include male (M) and female (F). The right panels are the distribution of genomic contexts at the DMR (centered) and surrounding regions. Genomic contexts include 1kb promoter, exon, intron, CpG island, predicted CTCF binding sites, public CTCF ChIP-seq peaks, enhancers, and repetitive sequences. The line chart on top of each panel is the average enrichment score for all the DMRs.

**Figure S2.**
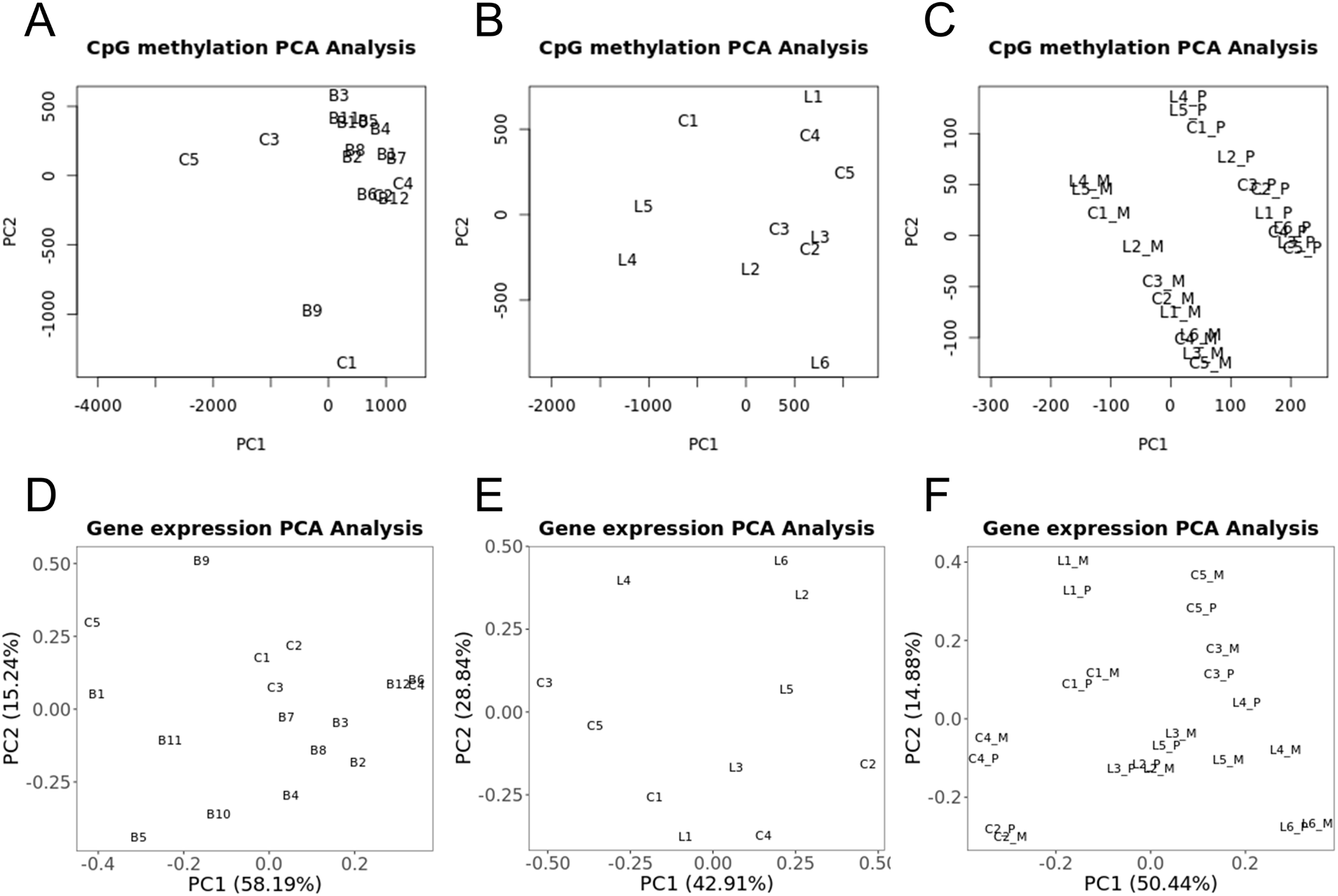
Principal component analysis (PCA) of DNA methylation and gene expression data. **(A)**, **(B)** and **(C)** DNA methylation PCA for human **(A)**, bovine **(B)**, and bovine allele-specific **(C)**. **(D)**, **(E)** and **(F)** Gene expression PCA for human **(D)**, bovine **(E)**, and bovine allele-specific **(F)**. Abbreviations for groups are control (C), BWS (B), and LOS (L). Sample numbers immediately follow group letters. Abbreviations for sub-groups are maternal allele (_M) and paternal allele (_P).

**Figure S3.**
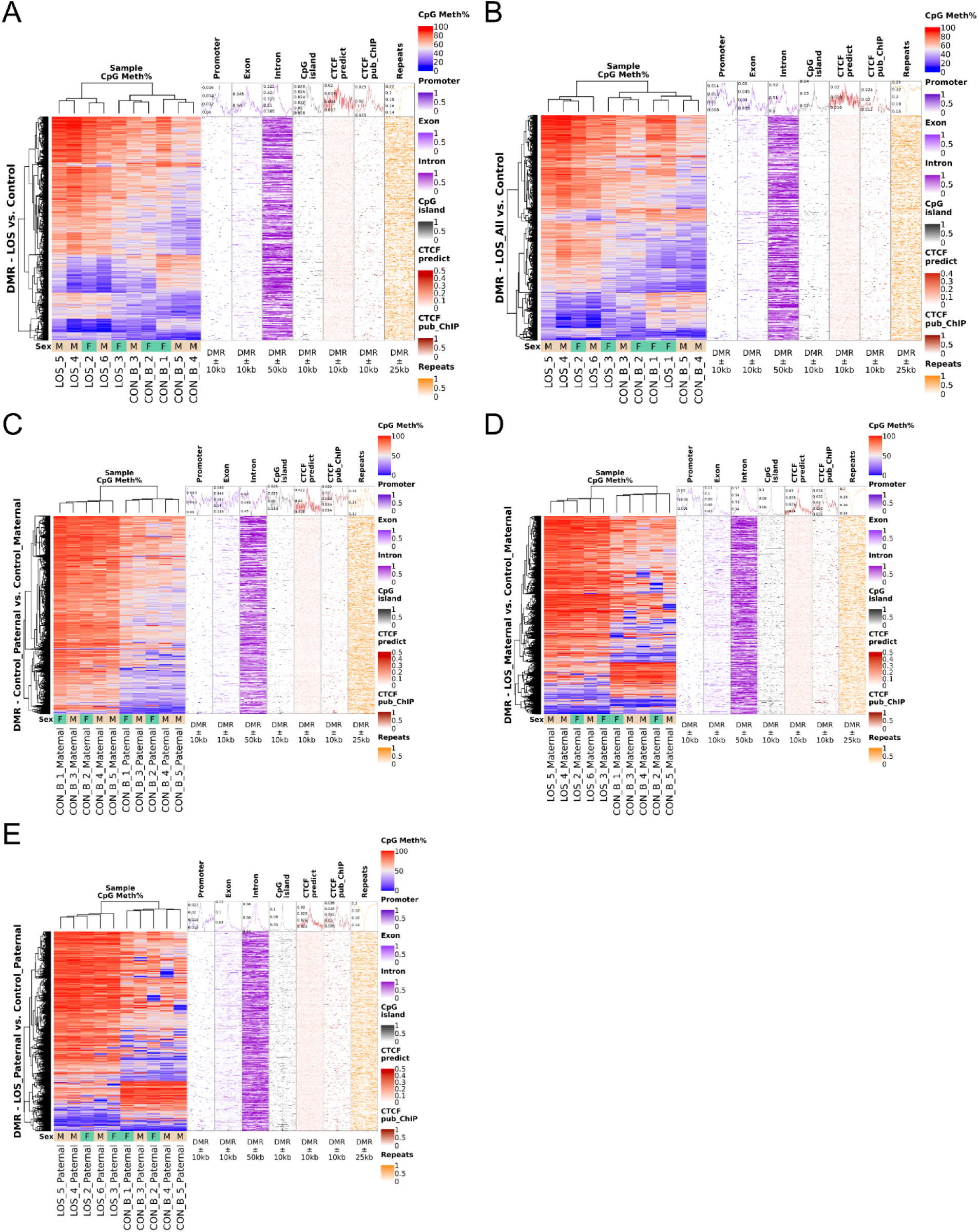
CpG methylation heatmap and genomic context enrichment for compared with control group. **(A),** LOS_All group compared with control group **(B)**, and allele-specific comparisons including Control_Paternal group compared with Control_Maternal group **(C)**, LOS_Maternal group compared with Control_Maternal group **(D)**, and LOS_Paternal group compared with Control_Paternal group **(E)**. The left panel is the heatmap with hierarchical clustering and sex annotation which include male (M) and female (F). The right panels are the distribution of genomic contexts at the DMR (centered) and surrounding regions. Genomic contexts include 1kb promoter, exon, intron, CpG island, predicted CTCF binding sites, public CTCF ChIP-seq peaks, and repetitive sequences. The line chart on top of each panel is the average enrichment score for all the DMRs.

**Figure S4.**
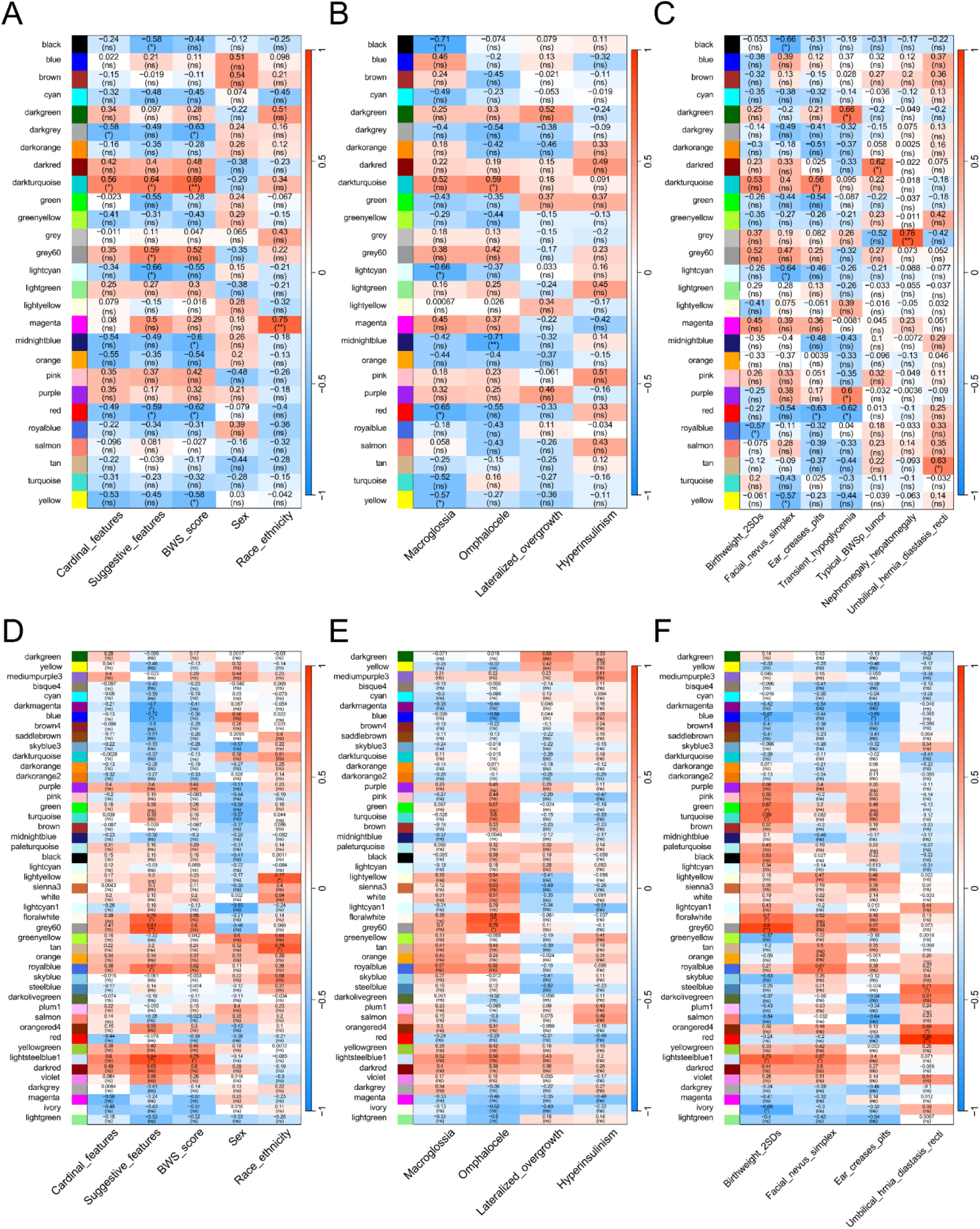
Identification of correlation network in BWS. (A-C) Correlation between cardinal features **(B)** and suggestive features **(C)** determined by weighted gene co-expression network analysis (WGCNA) for BWS subgroup 1 and control samples. **(D-F)** Correlation between gene expression profile in 47 modules and BWS diagnosis and scores **(D)**, BWS cardinal features **(E)** and suggestive features **(F)** determined for BWS subgroup 2 and control samples. Numbers in the heatmap represent correlation coefficient. Significance is shown as * (<0.05), ** (<0.01), and ns (≥0.05) in parentheses.

**Figure S5.**
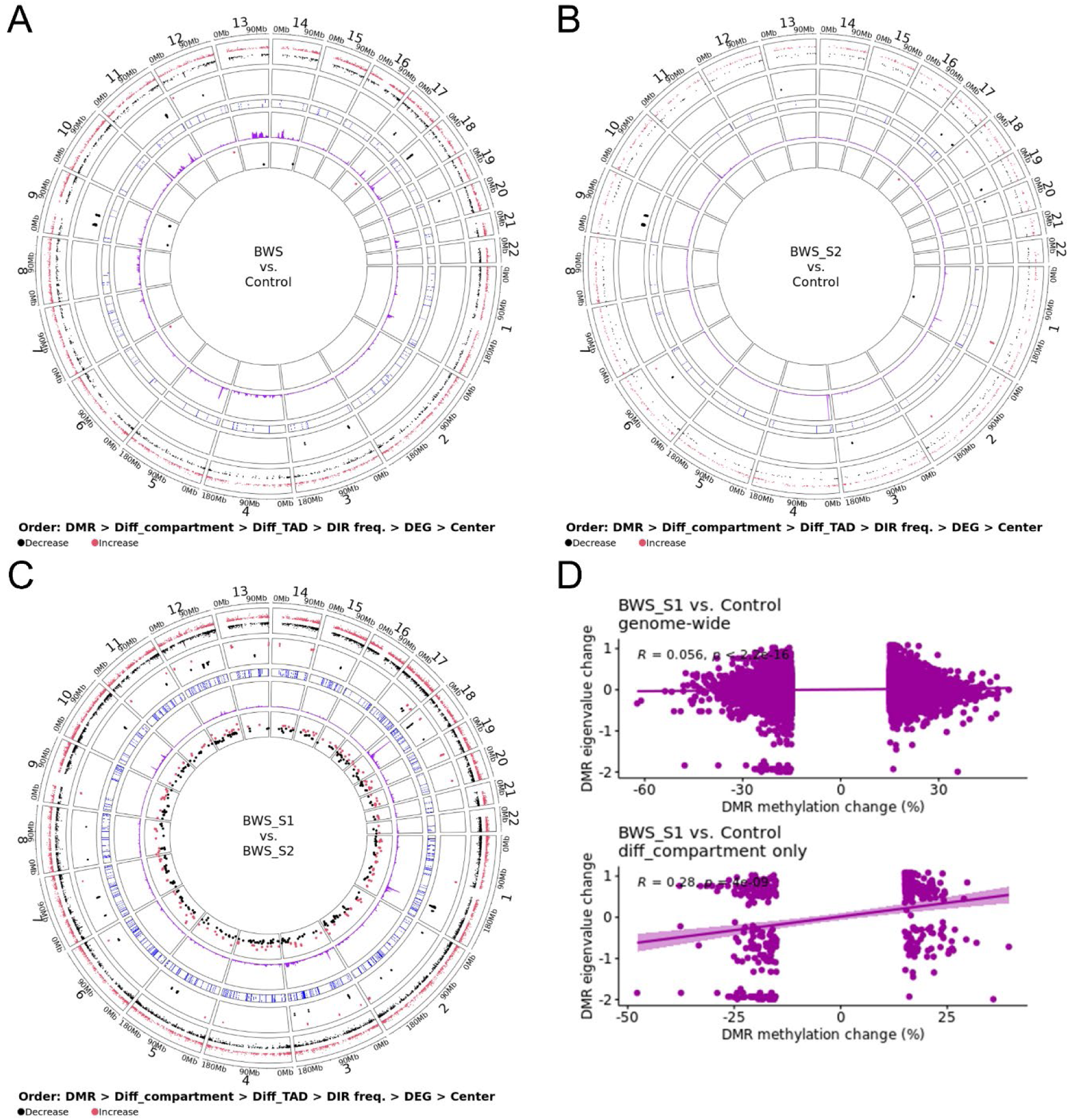
Genome-wide distribution of differentially methylated regions (DMR), differential compartments (diff_compartment), differential TAD (diff_TAD), differential interacting regions (DIR) frequency, and differentially expressed genes (DEG) identified in human group comparisons. Red dots indicate an increase in CpG methylation, eigenvalue (towards A compartment), and expression level, and black dots indicate decrease. The vertical location of dots indicates the level of changes. Mb = megabases. **(D)** Correlation between chromosome compartment changes (measured by wide correlations, and the bottom panel shows correlation at regions of differential compartment.

**Figure S6.**
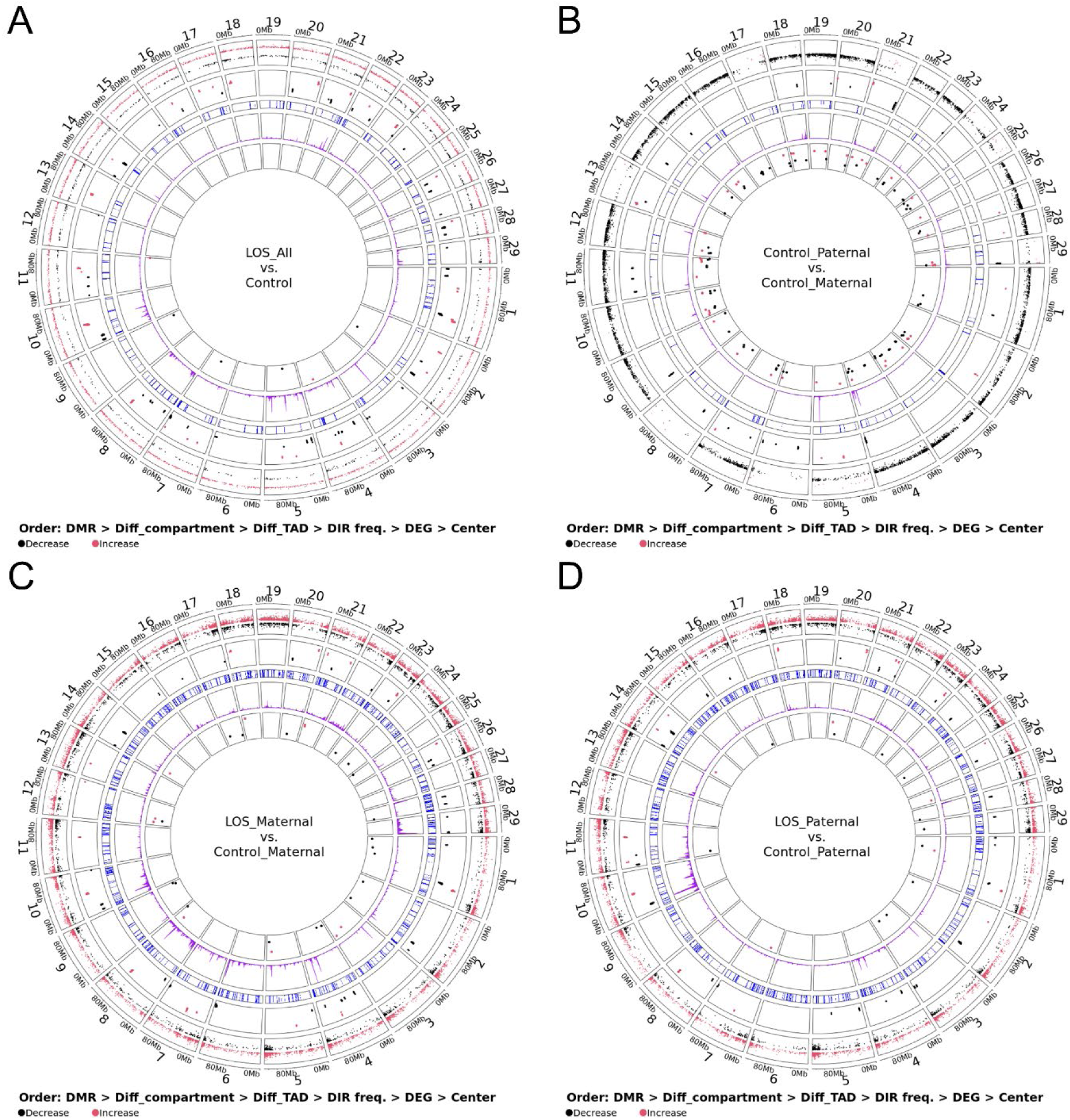
Genome-wide distribution of differentially methylated regions (DMR), differential compartments (diff_compartment), differential TAD (diff_TAD), differential interacting regions (DIR) frequency, and differentially expressed genes (DEG) identified in bovine group comparisons. Red dots indicate an increase in CpG methylation, eigenvalue (towards A compartment), and expression level, and black dots indicate decrease. The vertical location of dots indicates the level of changes. Mb = megabases.

**Figure S7.**
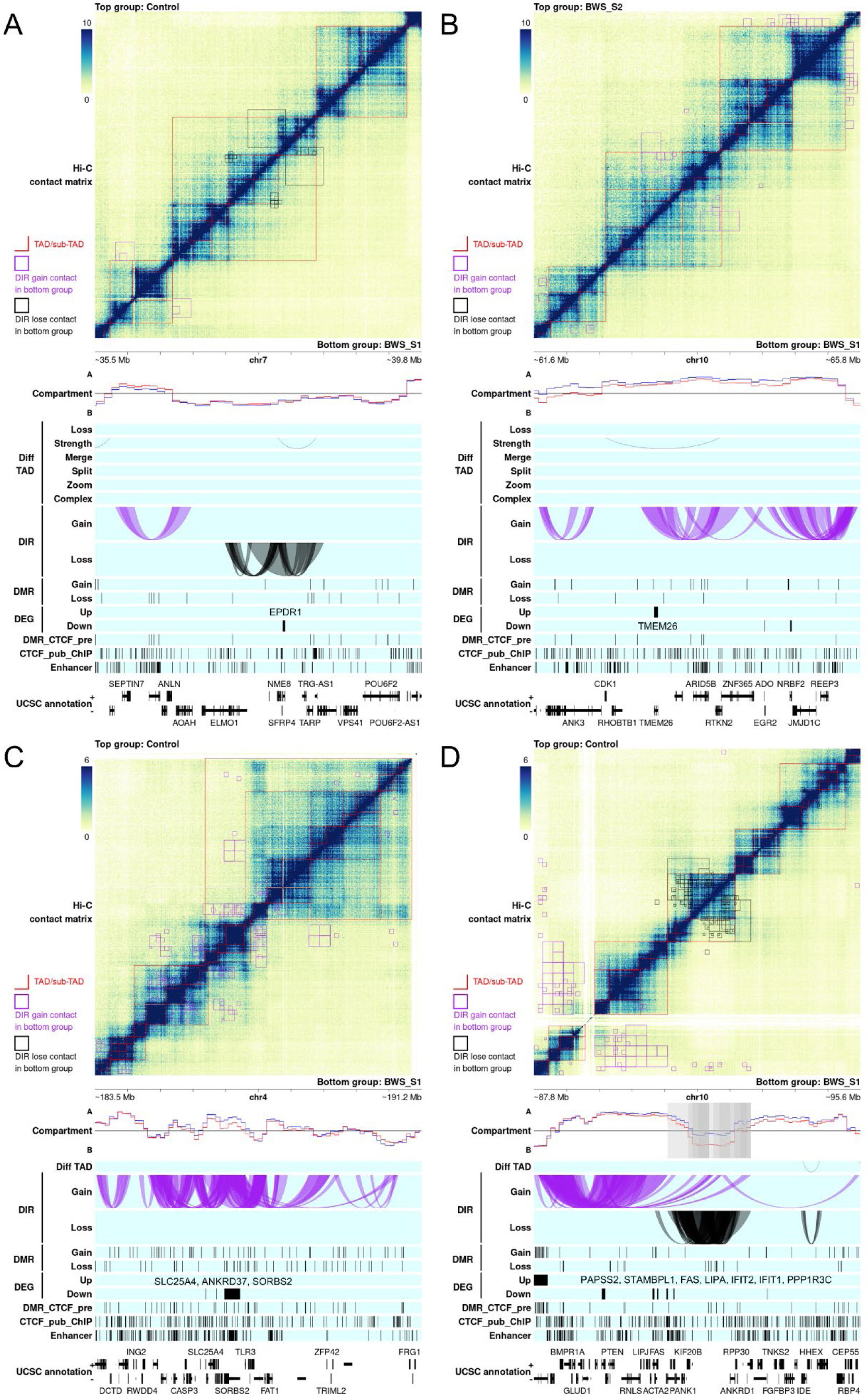
Examples of chromosome architecture alterations in human Beckwith-Wiedemann Syndrome (BWS). The top matrix plot shows the Hi-C contacts at 10kb resolution. A deeper blue color indicates higher physical contact frequency. Red lines mark the topologically associating domains (TADs). All the statistical analyses are conducted by comparing the bottom right group with the top left group. Tracks below the matrix show the location of different elements along this genomic region. The compartment track shows the A/B compartment represented by eigenvalue. The blue and red lines indicate the top left and bottom right group, respectively. The grey area marks significantly differential compartments between groups detected at different resolutions which may overlap and show deeper color. The diff_TAD tracks show the differential TADs in the second group. The DIR tracks show the differential interacting regions (DIR) that gain or lose contact. The DIRs are also marked in the matrix with black squares. The DMR tracks show differentially methylated regions (DMR) that gain or lose CpG methylation. The DEG tracks show up- or down-regulated differentially expressed genes (DEG). The DMR_CTCF_pre track shows predicted CTCF binding sites that overlap with DMRs. The CTCF_pub_ChIP track shows the public CTCF ChIP-seq peaks. The enhancer track shows common enhancer regions shared by at least 3 tissues/cell types related to BWS. The UCSC annotation tracks show the UCSC gene annotation.

**Figure S8.**
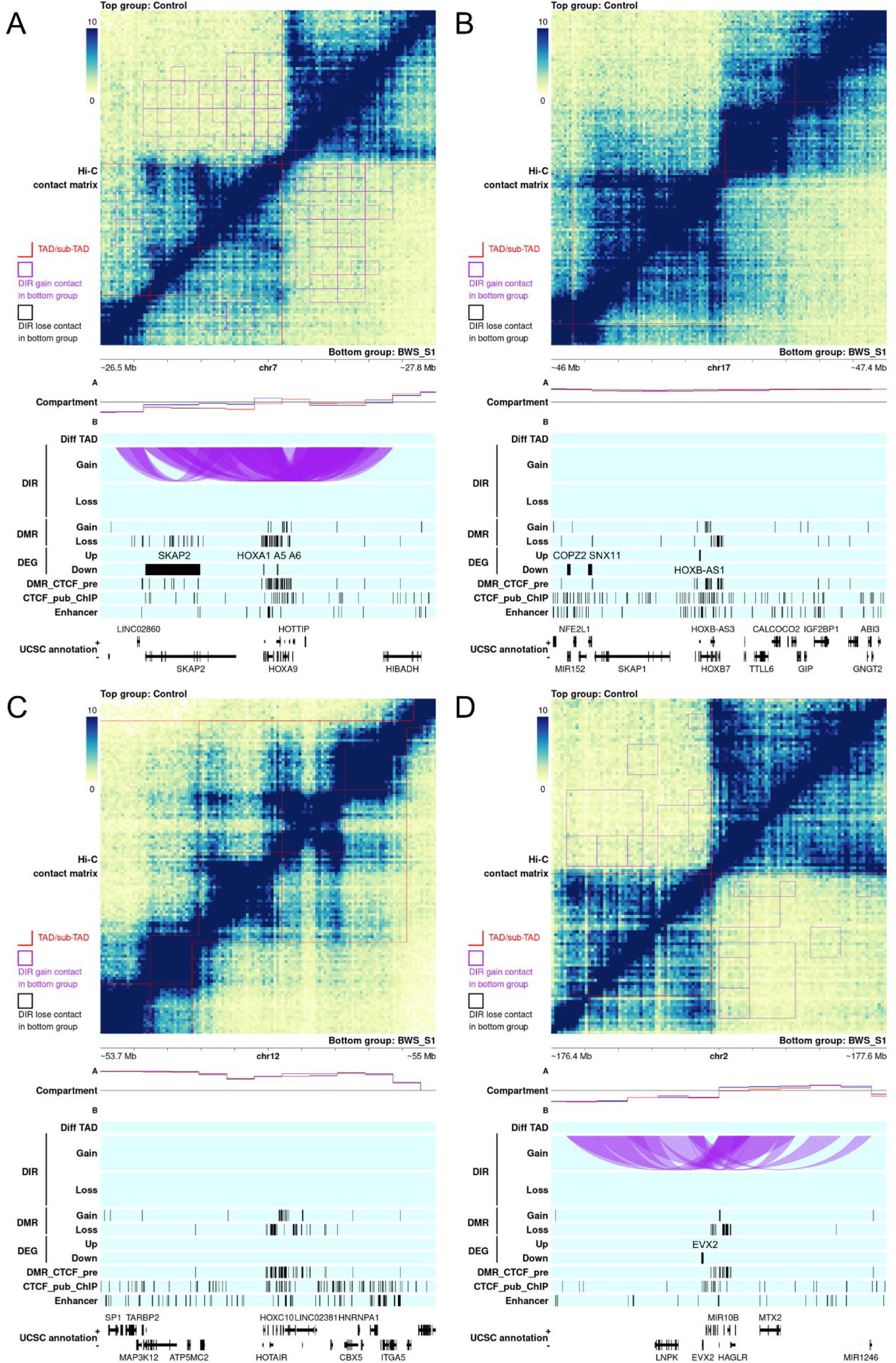
Chromosome architecture of *HOX* gene clusters in human Beckwith-Wiedemann Syndrome (BWS). *HOXA* **(A)**, *HOXB* **(B)**, *HOXC* **(C)**, and *HOXD* **(D)** clusters in human control and BWS subgroup 1. The top matrix plot shows the Hi-C contacts at 10kb resolution. A deeper blue color indicates higher physical contact frequency. Red lines mark the topologically associating domains (TADs). All the statistical analyses are conducted by comparing the bottom right group with the top left group. Tracks below the matrix show the location of different elements along this genomic region. The compartment track shows the A/B compartment represented by eigenvalue. The blue and red lines indicate the top left and bottom right group, respectively. The grey area marks significantly differential compartments between groups detected at different resolutions which may overlap and show deeper color. The diff_TAD tracks show the differential TADs in the second group. The DIR tracks show the differential interacting regions (DIR) that gain or lose contact. The DIRs are also marked in the matrix with black squares. The DMR tracks show differentially methylated regions (DMR) that gain or lose CpG methylation. The DEG tracks show up- or down-regulated differentially expressed genes (DEG). The DMR_CTCF_pre track shows predicted CTCF binding sites that overlap with DMRs. The CTCF_pub_ChIP track shows the public CTCF ChIP-seq peaks. The enhancer track shows common enhancer regions shared by at least 3 tissues/cell types related to BWS. The UCSC annotation tracks show the UCSC gene annotation.

**Figure S9.**
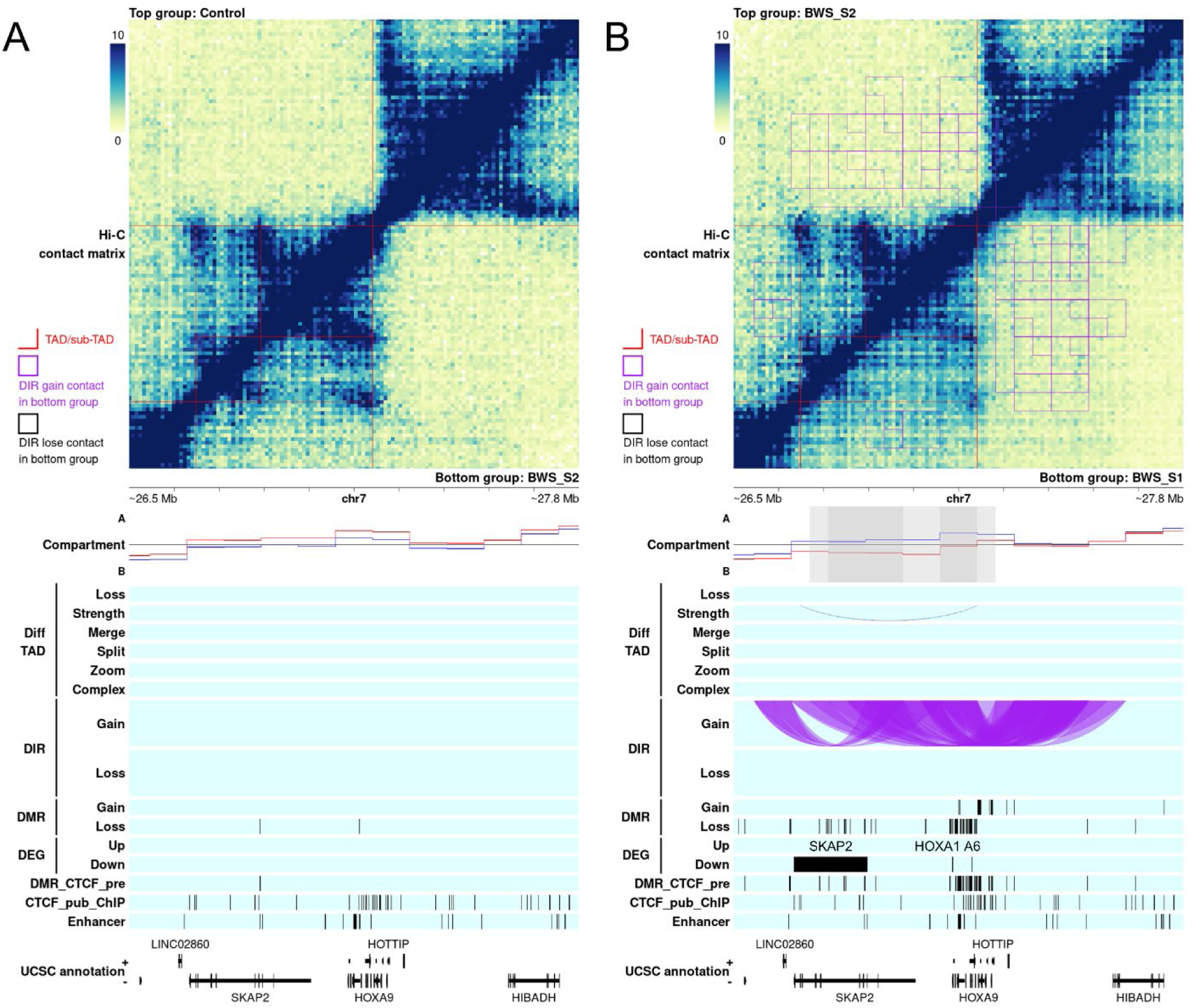
Chromosome architecture of *HOXA* cluster in human Beckwith-Wiedemann Syndrome (BWS). *HOXA* cluster for BWS subgroup 2 compared to control **(A)** and BWS subgroup 1 compared to subgroup 2 **(B)**. The top matrix plot shows the Hi-C contacts at 10kb resolution. A deeper blue color indicates higher physical contact frequency. Red lines mark the topologically associating domains (TADs). All the statistical analyses are conducted by comparing the bottom right group with the top left group. Tracks below the matrix show the location of different elements along this genomic region. The compartment track shows the A/B compartment represented by eigenvalue. The blue and red lines indicate the top left and bottom right group, respectively. The grey area marks significantly differential compartments between groups detected at different resolutions which may overlap and show deeper color. The diff_TAD tracks show the differential TADs in the second group. The DIR tracks show the differential interacting regions (DIR) that gain or lose contact. The DIRs are also marked in the matrix with black squares. The DMR tracks show differentially methylated regions (DMR) that gain or lose CpG methylation. The DEG tracks show up- or down-regulated differentially expressed genes (DEG). The DMR_CTCF_pre track shows predicted CTCF binding sites that overlap with DMRs. The CTCF_pub_ChIP track shows the public CTCF ChIP-seq peaks. The enhancer track shows common enhancer regions shared by at least 3 tissues/cell types related to BWS. The UCSC annotation tracks show the UCSC gene annotation.

**Figure S10.**
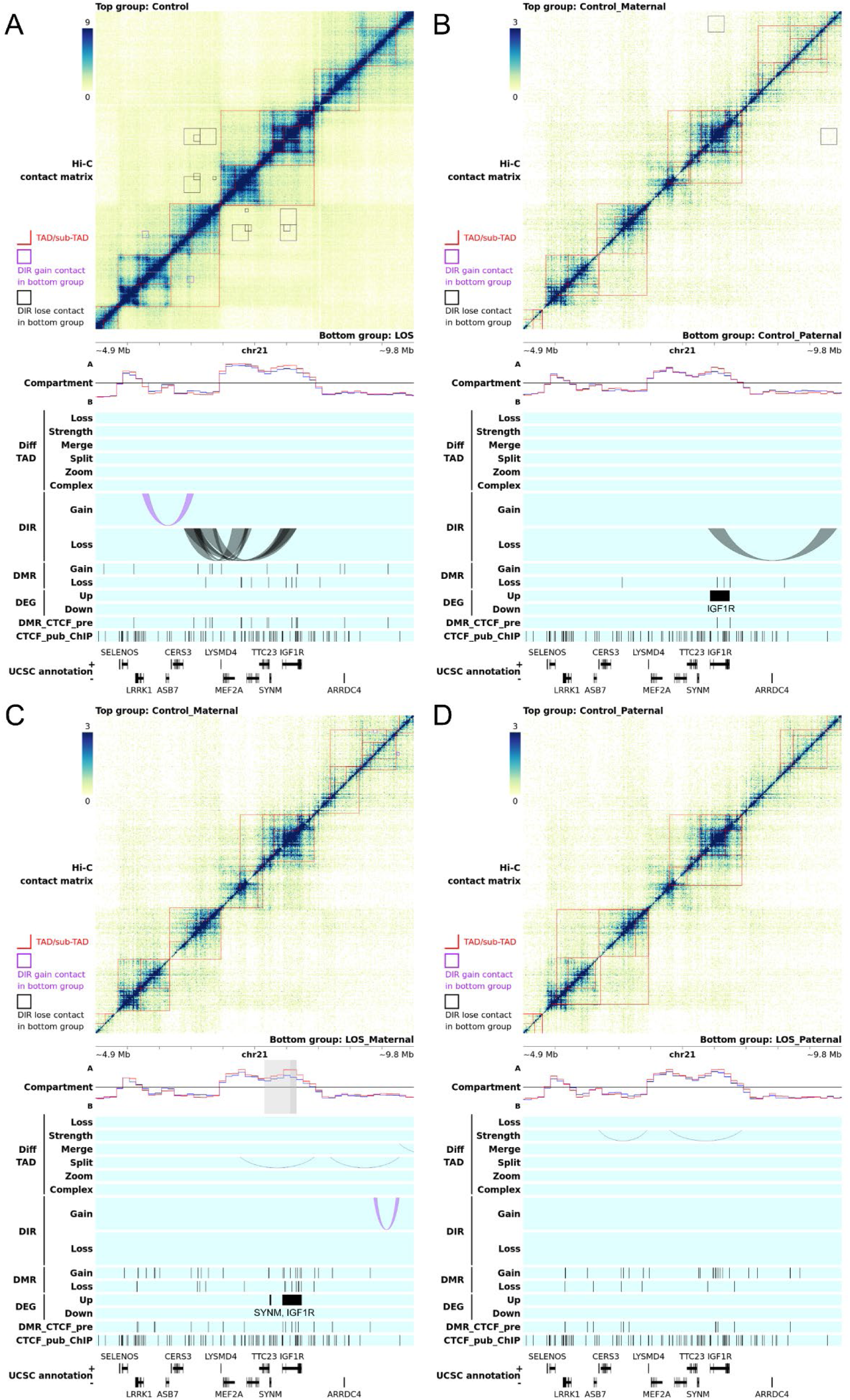
Examples of chromosome architecture alterations in bovine large offspring syndrome (LOS). The top matrix plot shows the Hi-C contacts at 10kb resolution. A deeper blue color indicates higher physical contact frequency. Red lines mark the topologically associating domains (TADs). All the statistical analyses are conducted by comparing the bottom right group with the top left group. Tracks below the matrix show the location of different elements along this genomic region. The compartment track shows the A/B compartment represented by eigenvalue. The blue and red lines indicate the top left and bottom right group, respectively. The grey area marks significantly differential compartments between groups detected at different resolutions which may overlap and show deeper color. The diff_TAD tracks show the differential TADs in the second group. The DIR tracks show the differential interacting regions (DIR) that gain or lose contact. The DIRs are also marked in the matrix with black squares. The DMR tracks show differentially methylated regions (DMR) that gain or lose CpG methylation. The DEG tracks show up- or down-regulated differentially expressed genes (DEG). The DMR_CTCF_pre track shows predicted CTCF binding sites that overlap with DMRs. The CTCF_pub_ChIP track shows the public CTCF ChIP-seq peaks. The UCSC annotation tracks show the UCSC gene annotation.

## Notes

### Competing Interest Statement

The authors have declared no competing interest.

### Author Declarations

Ethics committee/IRB of Childrens Hospital of Philadelphia gave ethical approval for this work

### Summary of Updates

The data were reanalyzed and the manuscript was rewritten for clarity

